# Healthcare workers’ prioritization of snake antivenoms for the treatment of snakebite envenoming: Perspectives from Ghana

**DOI:** 10.1101/2025.06.12.25329513

**Authors:** Eric Nyarko, Ebenezer Kwesi Ameho

**Affiliations:** Department of Statistics and Actuarial Science, College of Basic and Applied Sciences, University of Ghana, Legon, Accra, Ghana

**Keywords:** Best-worst scaling, Snakebites, Snakebite envenomation, Ghana, Sub-Saharan Africa, Neglected Tropical Diseases

## Abstract

**Introduction:** In low- and middle-income countries (LMICs) with tropical or subtropical climate and a high poverty rate the risk for snakebite envenoming (SBE) is high. Although patients ultimately receive antivenom treatment, healthcare workers are the primary end-users rather than the patients themselves. This study aimed to assess healthcare workers’ antivenom prioritization by providing quantitative evidence to guide policymakers to make better decisions to improve the procurement and supply of antivenoms, enhance the effectiveness of snakebite treatment, and improve patient care quality in health facilities in Ghana and other LMICs where SBE is common.

**Method:** We collected data by conducting an interview-based survey using questionnaires. We randomly selected 203 healthcare workers from the Kwahu Afram Plains North and South districts in the Eastern Region of Ghana in August 2023. We used the best-worst scaling experiment design method to assess healthcare workers prioritization of snake antivenoms available for use in sub-Saharan Africa.

**Result:** Among the antivenoms available for use in sub-Saharan Africa, participants highly prioritized Inoserp Pan-Africa polyvalent antivenom. Snake Venom Antiserum -PanAfrica is also commonly prioritized, followed by ASNA antivenom D, ASNA antivenom C, Snake Venom Antiserum African - 10, Anti Snake Venom Serum Pan Africa – 10, and Fav-Afrique. However, some antivenoms are least commonly prioritized, such as SAIMR Echis, Combipack of Snake Venom Antiserum, Anti Snake Venom Serum Central Africa -6, Anti-Snake Venom Serum Central Africa, Snake Venom Antiserum Echiven Plus, Antivipmyn-Africa, Menaven, Snake Venom Antitoxin, Snake Venom Antiserum (Echiven), Anti Snake Venom Serum Monovalent Echis ocellatus, EchiTAbG, and Snake venom antiserum Echis ocellatus (VINS-Echis). In comparison to other antivenoms, Inoserp Pan-Africa is more frequently prioritized. At the same time, Snake Venom Antiserum -PanAfrica is less frequently prioritized but still more likely than other options. EchiTabPlus (ET-Plus) is more likely to be prioritized than other antivenom options.

**Conclusion:** Our findings offer valuable insights to guide policy discussion on available antivenoms in treating SBE. There is an urgent need to implement regulations on antivenom products, improve procurement and supply, offer ongoing education, and provide training to healthcare workers to combat the burden of SBE.

**Author summary:** Healthcare workers are the primary users of snake antivenoms rather than the patients who receive them. Therefore, their antivenom prioritization can help policymakers to make better decisions to improve the procurement and supply of quality antivenoms, enhance the effectiveness of snakebite treatment, and improve patient care quality in health facilities in Ghana and other low- and middle-income countries where snakebite envenoming is prevalent. To this end, we conducted a study to assess healthcare workers’ antivenom prioritization and provide policymakers with quantitative evidence to guide decision-making. Using an interview-based questionnaire, we surveyed 203 healthcare workers in the Kwahu Afram Plains North and South districts in Ghana’s Eastern Region in August 2023. The best-worst scaling experimental design method was used to assess participants’ prioritization of different snake antivenom products available in sub-Saharan Africa. Most participants were female, aged 18 to 30 years old with 1-5 years of work experience. Many participants had received snakebite training and mostly lived in rural areas. The majority reported that farmers were the most commonly bitten population during the rainy season, in their farms or bush, between 9 am to 12 noon. Among the snake antivenom products in sub-Saharan Africa, polyvalent antivenoms were highly prioritized over monovalent ones. Inoserp Pan-Africa was the most frequently prioritized, followed by Snake Venom Antiserum -PanAfrica (Premium-A) and EchiTabPlus. Our findings provide valuable insights to guide policy discussions on available antivenoms in treating snakebite envenoming. We urge policymakers to implement regulations on antivenom products, improve quality antivenom procurement and supply, provide ongoing education, and offer training to healthcare workers to combat the burden of snakebite envenoming.

## 1. Introduction

Snakebite envenoming (SBE) is a severe and widespread condition primarily affecting marginalized communities in low- and middle-income countries (LMICs), such as rural villagers, agricultural workers, nomadic farmers, pregnant women, young males, and working children, who often have limited access to education and healthcare [1, 2, 3]. SBE is a medical emergency when snake venom is injected into the body through a bite or sprayed onto mucosal surfaces or eyes, causing severe bleeding, paralysis, kidney injury, muscle damage, and other local tissue damage [4, 2]. Depending on the venom’s toxicity, SBE can lead to permanent disability and death if left untreated [5].

Since 2017, SBE has been classified as a Category A Neglected Tropical Disease (NTD) by the World Health Organization (WHO) [1], and has launched a global strategy to prevent and control SBE, aiming to reduce deaths and disabilities by 50% and deliver three million effective treatments annually by 2030 [6]. According to estimates, 5.4 million people worldwide are bitten by snakes yearly, with 2.7 million experiencing envenoming [2]. SBE is responsible for approximately 83,000-138,000 deaths per annum, and approximately 400,000 people per annum suffer from disabilities such as amputations, scarring leading to impaired limb function, and post-traumatic stress disorder. In sub-Saharan Africa (SSA) alone, SBE is estimated to affect 435,000-500,000 people annually, with 20,000–32,000 deaths [7]. The hospital visit rate of snakebite victims in rural SSA alone was estimated at 56.4/100,000, and mortality was at 1.35/100,000 inhabitants [3]. In Ghana, rural health facility data put the incidence of snakebite at 35/100,000 inhabitants per annum, with up to 11% case fatalities [8, 9].

Among, the approximately 110 Africa venomous snake species, WHO considers 24 species from four genera (Bitis, Dendroaspis, Echis, and Naja) as having the highest (Category 1) medical importance (i.e., venomous snakes that are most encountered and pose the greatest potential threat to human life and wellbeing) in SSA. A further 24 species are of secondary (Category 2) medical importance (i.e., either because they are known to be highly venomous, but are either less frequently associated with serious snakebites, or have little epidemiological data available) [2]. Bites from these categories of snake species are often treated using heterologous animal plasma-derived immunoglobulin preparations, also known as antivenoms (here defined as purified antibodies against snakebite envenoming). Although antivenom products exist and are currently available with active pharmaceutical ingredient (API) (here defined as the specific antibody component that is produced through their production process) as either intact (whole) immunoglobulins (IgG), or as divalent fragment antibodies (F(ab’)2) or, more rarely, monovalent fragment antibodies (Fab)) [1, 2], they are mainly classified as ineffective, untested, and weakly regulated in SSA [1]. This broadly negative narrative has given rise to substantial scepticism and concern over the usefulness of antivenoms [2]. Recent studies have investigated the immunomodulatory effects of traditional medicinal compounds [10, 11], suggesting potential complementary therapies that may influence antivenom prioritization.

However, efficient antivenom treatment is crucial in the first hours after a snakebite [4]. Unfortunately, antivenom availability and accessibility remain distant possibilities for most regions where SBE is endemic [12]. Instead of being available at rural health facilities, antivenoms are often only accessible in large referral hospitals [13]. In Ghana, it was reported that antivenoms supplied to designated health facilities were insufficient to meet the demand [14]. Only about 20% of healthcare workers in Uganda, Zambia, and Kenya reported available antivenom stock in 0–34% of facilities across the sectors and countries [15]. Antivenom was available in 45% of public and less than 20% of private facilities in Kenya [16].

Currently, most antivenoms available in SSA are broad-spectrum, Pan-African polyvalent products designed for WHO Category 1 Bitis, Dendroaspis, Echis, and Naja species. While some West African countries have monovalent antivenoms for bites by the Category 1 carpet vipers *Echis romani/Echis ocellatus*, there are limited quantities available for less common bites by the Category 2 colubrid *Dispholidus typus* [2]. In Ghana, the Food and Drug Authority (FDA) provides guidelines for antivenom registration [17]. All the three registered antivenoms (i.e., Anti Snake Venom Serum Pan Africa – 10, Snake Venom Antiserum -PanAfrica (Premium-A), Snake Venom Antiserum African - 10 (Afriven 10)) [18] are polyvalent and of equine origin.

Although the availability of antivenoms at health facilities involves various processes, including development, manufacture, registration, pricing, procurement, supply, prescribing, and dispensing [4], early diagnosis of symptoms and signs of snakebite envenoming by healthcare workers is crucial for appropriate antivenom administration [2]. Therefore, understanding healthcare workers’ antivenom prioritization is essential to provide strategies to improve patient safety and treatment efficacy in Ghana and other LMICs where SBE is common. The available research primarily focuses on the burden of snakebite, control, and management, estimated cost to meet, projected antivenom need, availability, accessibility, management, and use of antivenoms [3, 19, 20, 21, 4, 22, 23, 14, 24], but there is lack of data on healthcare workers’ antivenom prioritization. In light of WHO’s strategy for the prevention and control of SBE, it is necessary to improve the information on snake antivenoms available for use in sub-Saharan Africa to have a more robust platform to build strategies to combat this NTD. The present study aims to address this gap by assessing healthcare workers’ antivenom prioritization. The study was conducted in the Eastern Region of Ghana and focused on two districts, namely Kwahu Afram Plains North District and Kwahu Afram Plains South District. Using the best-worst scaling (BWS) methodology, we provide a detailed quantification of healthcare workers’ antivenom prioritization. Policymakers and healthcare providers can leverage this information to enhance the procurement and supply of antivenoms, improve the efficacy of snakebite treatment, and elevate patient safety and care quality in health facilities in Ghana and SSA, where snakebite envenoming is a significant concern.

## 2. Methods

### 2.1. Ethical approval and consent to participate

The study followed ethical guidelines and regulations. Ethical clearance was obtained from the Ethics Committee of the College of Basic and Applied Sciences, University of Ghana (Reference No: ECBAS 065/22-23). Before collecting data, written informed consent was obtained from all respondents after explaining the purpose of the study. It was emphasized that participation in the study was voluntary, and respondents had the choice to decline. Respondents’ confidentiality and anonymity were safeguarded using unique identifiers.

### 2.2. Study site

The study was conducted in the Eastern Region (representing 9.5% of Ghana’s population [25]) of Ghana and focused on two districts, namely Kwahu Afram Plains North District (KAPND) and Kwahu Afram Plains South District (KAPSD). These districts were selected based on reported cases of snake bites [26] and consultation with primary healthcare personnel. The KAPND is bordered to the south by KAPSD, the Volta River to the east, and two districts in the Ashanti Region (Sekyere East and Asante-Akim) to the west. KAPND, which covers an area of 2,570 square kilometers, has a population density of 25.9 persons per square kilometer and a population of 66,555, representing 2.3% of the regional population. The male population is 53.4%, the female population is 46.6%, and the average household size is 3.7 [27, 25]. KAPSD is bordered north by KAPND, Kwahu South to the south, the Volta River to the east, and two districts in the Ashanti region (Sekyere East and Ashanti-Akim) to the west. KAPSD covers an area of 3,051 square kilometers, has a population density of 24.3 persons per square kilometer, and a population of 74,002, representing 2.5% of the regional population. The male population is 53.3%, the female population is 46.7%, and the average household size is 3.8 [27, 25].

Both districts are in the savannah vegetation zone, with short deciduous fire-resistant trees and varying grass heights. The rainfall occurs in June and October (the first rainy season starts from May to June and the second from September to October), with the mean annual rainfall ranging from 1,150mm to 1,650mm.

The dry seasons start between November and late February, with high temperatures ranging from 36.6°C to 36.8°C in February and March, respectively, and low temperatures between 19.1°C and 20.1°C in December and January. The Afram and Volta rivers are the primary drainage sources in both districts and are utilized throughout the year for farming, fishing, and household activities [28, 27].

### 2.3. Sample size and data collection

To conduct this study, we surveyed healthcare workers in the KAPND and KAPSD districts of the Eastern Region of Ghana in August 2023. Data was collected from a simple random sample of 203 out of 698 healthcare workers, including community health nurses, physician assistants/medical doctors, and certificate/enrolled/general nurses in twelve healthcare facilities. The workers were contacted during working hours in the various health facilities. It is worth noting that some of the healthcare workers contacted may not administer antivenoms but involved in the management of snakebites. A prior study in Ghana indicated that medical doctors, registered nurses, physician assistants, community health nurses, and pharmacy technicians were the primary healthcare professionals involved in managing snakebites [29]. The minimum number of required respondents (N) is 81, based on a sample size calculation [30] and the available scenarios (*l=*19) using the formula *N=n/l*. Here, *n* ≥ *r*^2^*s*/*va*^2^, where r = Φ^−1^(1 − *α*/2) represents the inverse cumulative distribution function of a standard normal distribution reported at (1 − *α*/2), *v* is the true choice proportion of the study population obtained under the assumption of equal choice probabilities, *a* is the allowable deviation as a percentage between the estimate *v̂* and *v*, *s* = 1 − *v*, *α* is the level of 95% confidence in the estimations such that Pr (|*v*^ − *v*| ≤ *av*) ≥ α for any given sample *n* under the standard normal distribution. The anticipated sample size of 203 respondents was enough, and all 203 distributed questionnaires were included in the final analysis because of no missing data. This was made possible because enumerators were available to assist and guide respondents in completing the survey instrument.

### 2.4. Best-worst scaling

In various fields such as healthcare, conflict resolution, environmental sustainability, and consumer research, the BWS approach is used to elicit preferences for various goods and services [31, 32, 33, 34, 35, 36]. Respondents are asked to choose the best (or most desired/preferred) and worst (or least desired/preferred) options from a list of at least three attributes within a BWS scenario or choice set. BWS method enables evaluation of the relative importance of attributes across multiple scenarios. This method requires more cognitive work and involvement than typical rating scale tasks, which might help respondents complete a decision task [37]. Earlier works have also highlighted that BWS [38, 39] is more appealing than discrete choice experiment design approach [40].

In this study, we utilized BWS Case 1 [41]. We started with an initial list of 28 attributes related to snake antivenoms based on a literature review [20, 1, 4]. Notably, although there was some overlap, 6, 2, and 20 of the antivenoms appeared in [1], [4], and [20], respectively. Through focus group discussions with healthcare professionals, we validated and narrowed this list down to twenty plausible snake antivenoms. Notwithstanding, a literature scrutiny was carried out using ScienceDirect [https://www.sciencedirect.com/#life-sciences] PubMed [https://pubmed.ncbi.nlm.nih.gov] and Google Scholar [https://scholar.google.com] databases to unearth published works on snake antivenoms that have been used or available/recommended for use in the management and treatment of SBE in Ghana. Although may not be exhaustive, the search revealed 11 of such antivenoms, of which only three are registered with the FDA (Table 1). Table 1 provides information on the snake antivenom products available for treating snakebite envenoming in SSA, including the brand name, the facilities producing them, the country of production, and the snake species against which the antivenom is active.

**Table 1.**
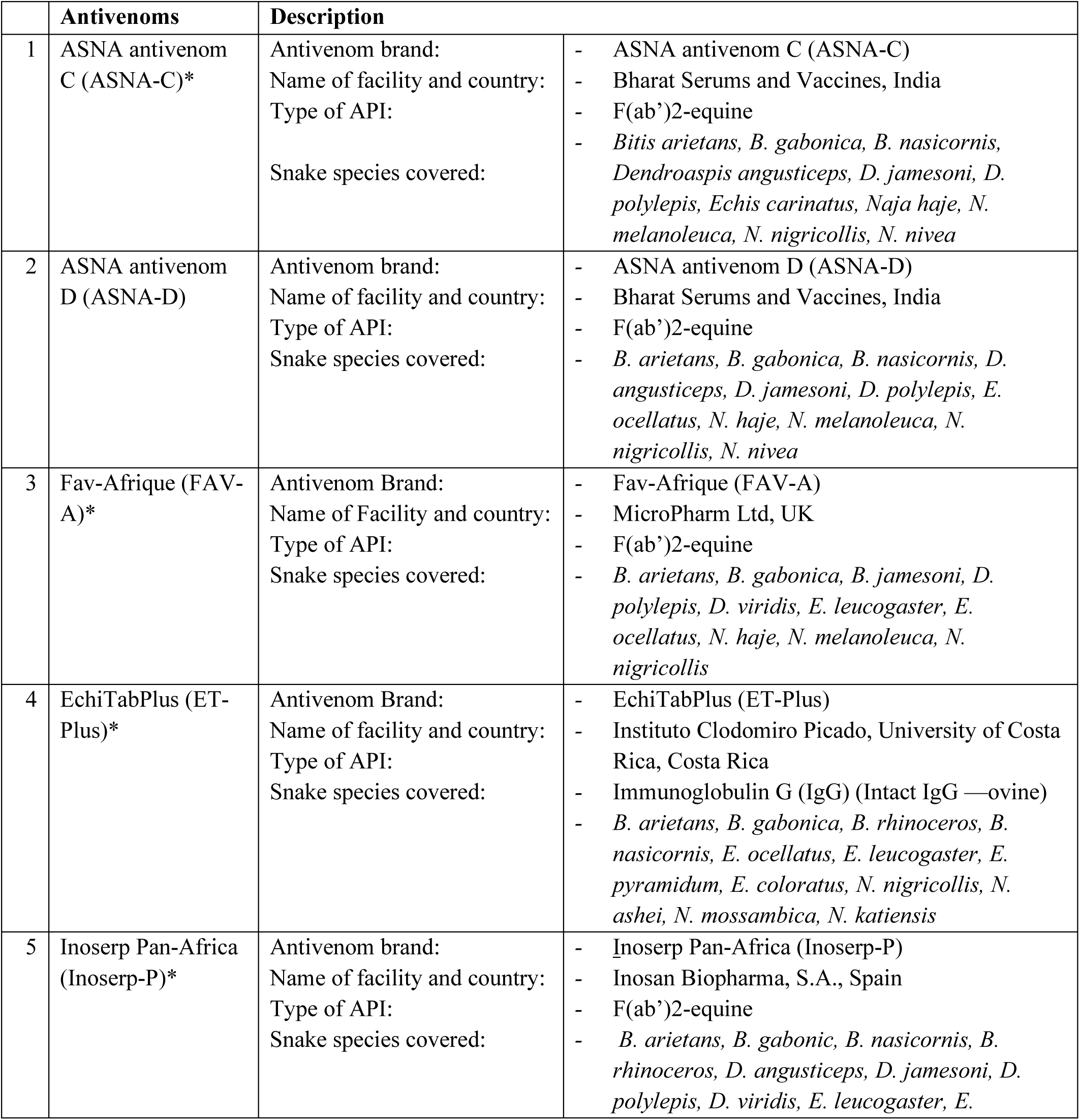
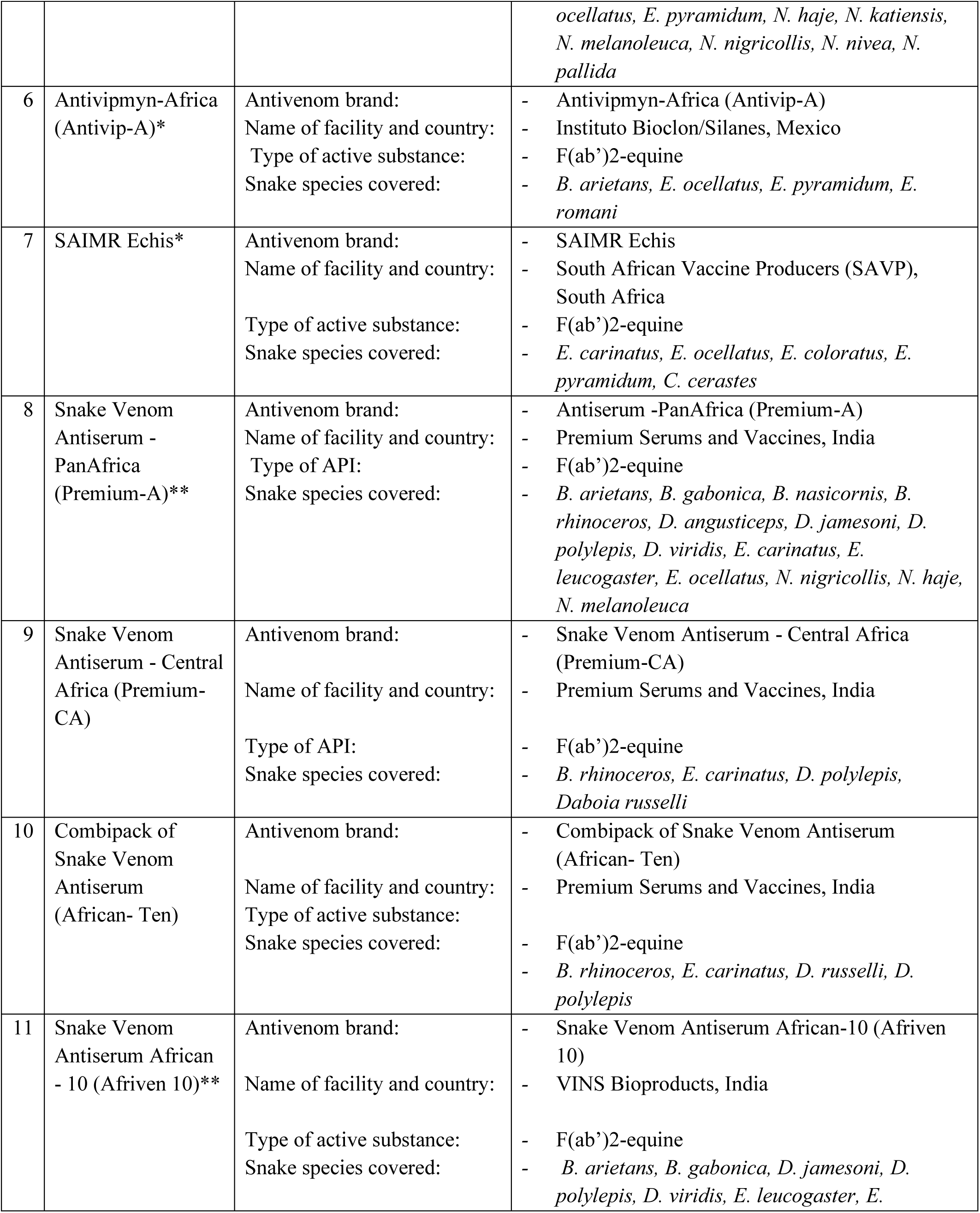
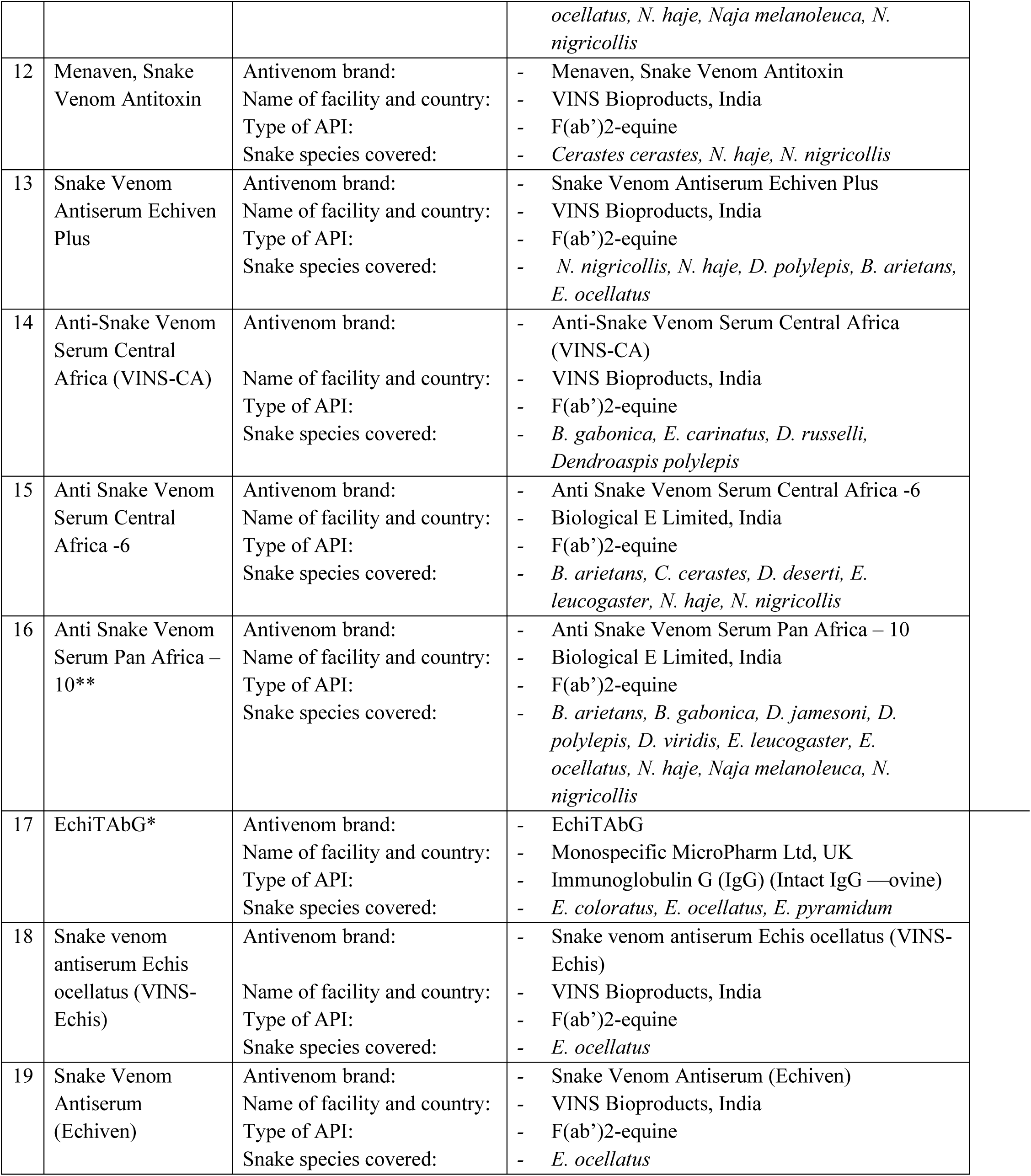
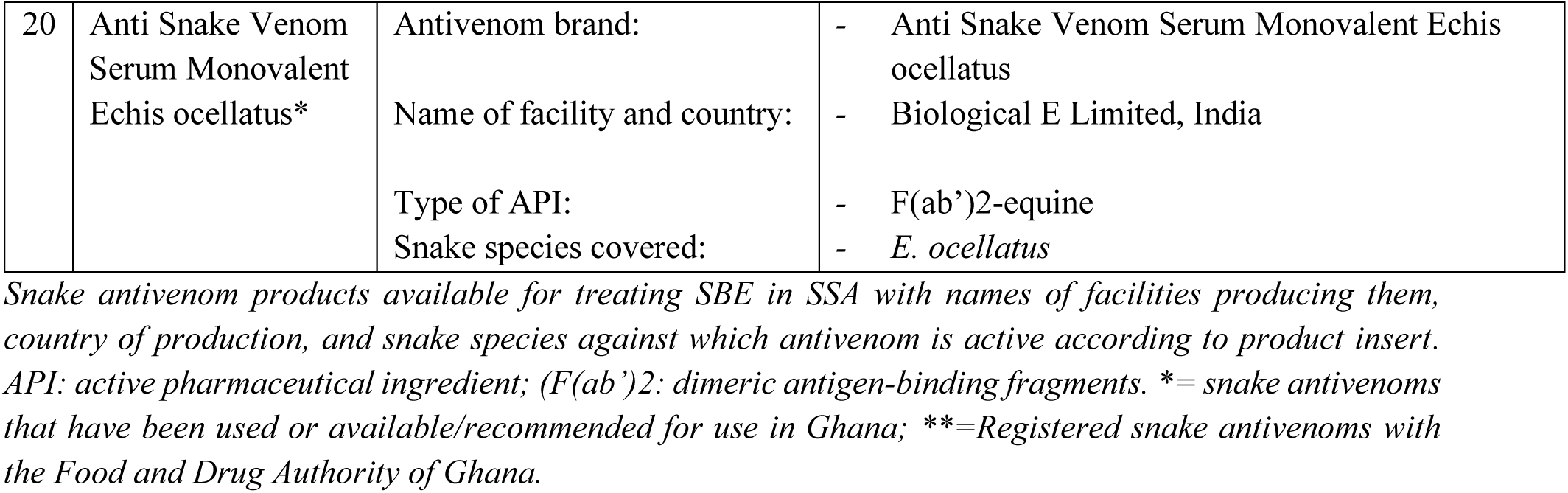
Snake antivenoms available for treating snakebite envenoming.

A statistical experiment block design [42] was used to randomly assign the snake antivenoms products in the BWS. We created 95 BWS scenarios, each containing four snake antivenoms. To make it easier for participants, we divided the 95 BWS scenarios into five equal blocks of 19 scenarios each [34]. Each participant was randomly assigned to one block with 19 BWS scenarios/questionnaires, each with four antivenoms. Through an interviewer-administered questionnaire, participants were asked to choose which antivenom they prioritize most and the least by writing out their responses using paper and pencil. We calculated the relative importance of each snake antivenom based on the frequency of choices made by participants. We collected demographic data from participants and asked them some questions about their experience or background before starting the survey. We also pilot tested the questionnaire with ten healthcare workers to ensure that respondents understood the definition of the snake antivenom products and could make trade-offs between them. Participants were verbally informed about the snake antivenom profiles and asked to inform the interviewer if any part of the survey needed clarification or was easier to answer. Participants found the survey questionnaire to be transparent and easy to answer. Although some participants believed that some snake antivenoms might not be available in the health facilities, none suggested any other antivenom for inclusion in the study. Fig. 1 shows a completed example BWS scenario.

**Fig. 1.**
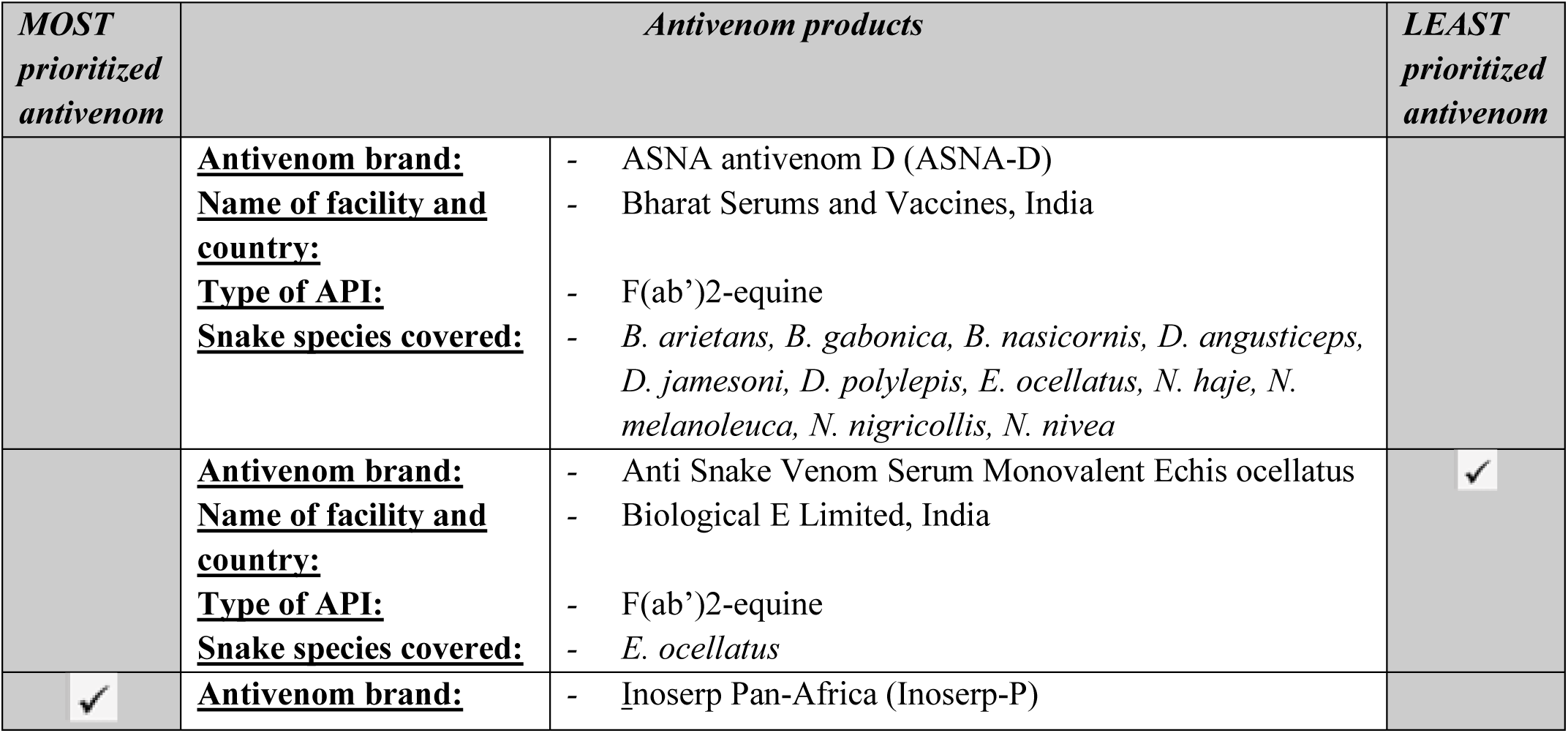
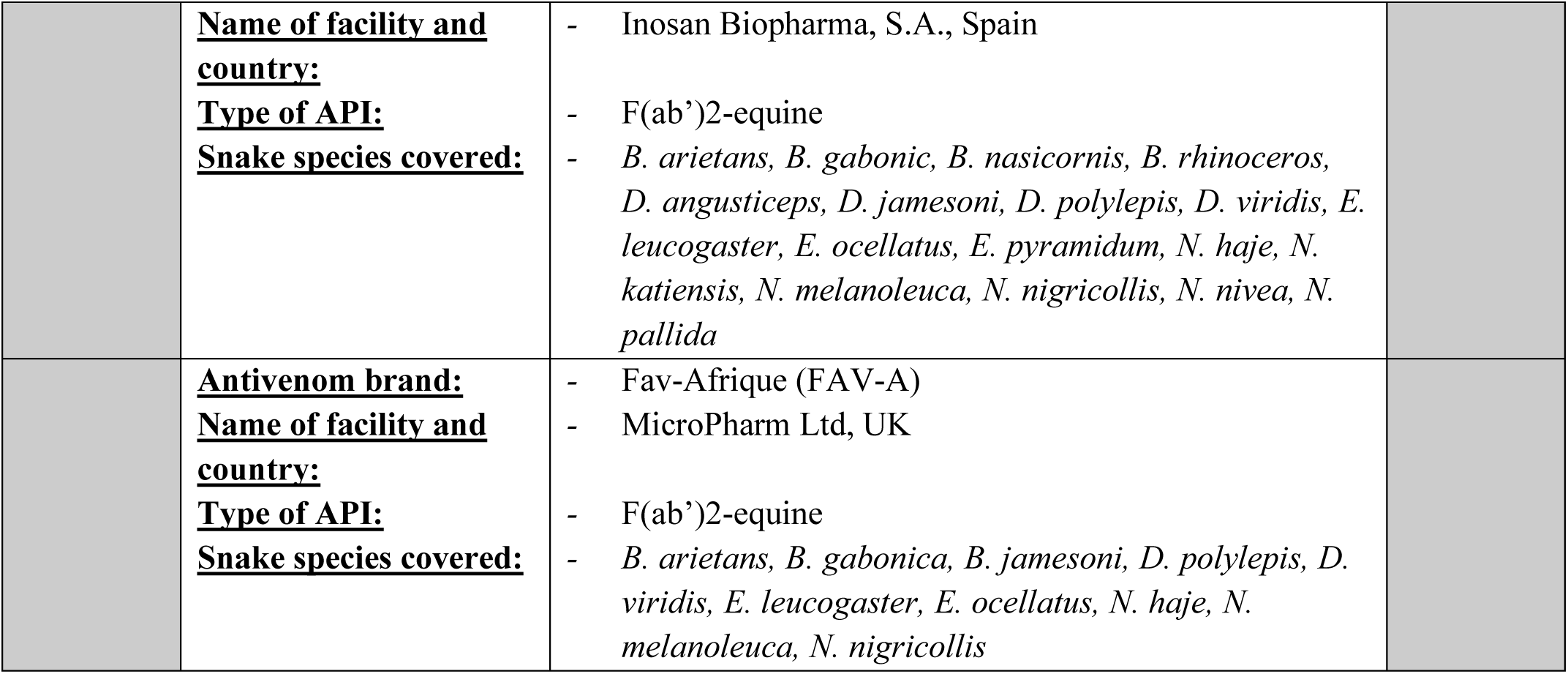
A sample completed BWS scenario presented to respondents. *The set of BWS scenarios generated for this study contain four antivenoms each, and the sample completed scenario represents BWS scenario number 1 in the first block and contains antivenoms labelled as 2, 20, 5, and 3*.

## 3. Statistical analysis

### 3.1. Maximum difference model

In this study, healthcare workers were asked to prioritize different antivenoms based on several criteria, including the brand name, the facilities producing them, the country of origin, and the specific snake species for which the antivenom is effective. Participants were instructed to identify which antivenom they would prioritize the most and which they would prioritize the least from a provided set of scenarios (Figure 1, for example). Repeated choices by participants from the sets of unordered BWS scenarios reveal the trade-offs healthcare workers are willing to make between the antivenoms. This choice is modeled as a function of the antivenoms using random utility theory (RUT) [43, 44, 45]. RUT is based on the idea that people (healthcare professionals) will make their decisions on the attributes of a product (antivenom) as well as a certain amount of chance (a random component). The random component results from either the respondent’s antivenom prioritization being somewhat random or the researcher does not have the complete set of information available to the respondent [46]. By using a model with a random component, we can estimate how often one snake antivenom might be chosen or prioritized over another. Here, the frequency of choices made by participants are used as a metric to compare the relative importance of different snake antivenoms. We assume that each choice has an underlying value or utility to respondents. To estimate these utilities, we use the maximum difference model [35, 45] to analyze the data. Statistical significance was observed with a 95% confidence interval (CI) greater or less than zero. A parameter estimate is considered significant if the 95% CI is greater or less than zero. If the 95% CI overlaps, the parameter estimate is not significantly different. A significant positive/negative parameter estimate indicates a prioritization/trade-off for a particular snake antivenom product. In the absence of CIs, statistical significance is considered if the significant probability (*p*-value) is either less than or equal to 0.001, 0.01, and 0.05. All statistical analysis was conducted using JMP Pro (Version 16.0). It is worth noting that the utility estimates (UE) quantify the perceived importance of the matching level of the effect. Higher values indicate that the effect’s level is deemed more important. The standard errors (SE) measure the uncertainty surrounding the parameter estimate for each antivenom. The better the model fits the data, the smaller the SE of the parameter estimate. The greatest utility difference (GUD) is the maximum change in utility that can be achieved from a specific antivenom based on the plausible antivenoms in the BWS study.

## 4. Results

### 4.1. Demographic description

In this study, 203 healthcare workers participated, including community health nurses, physician assistants/medical doctors, and certificate/enrolled/general nurses. The analysis of variance revealed 2 degrees of freedom, a sum of squares of 0.620, a mean square of 0.310, an F-ratio of 0.465, and a p-value of 0.628, indicating no significant difference in mean responses by professional qualifications. Table 2 offers an overview of these healthcare workers. Out of the participants, 78 (38.4%) worked in community-based health planning and services (CHPS) compounds, 100 (49.3%) worked in health centres/clinics, and 25 (12.3%) worked in hospitals (Table 2). Of the 203 respondents, 141 (69.5%) were female and aged between 18 and 30 years old, while 62 (30.5%) were male. Most respondents had 1-5 years of working experience 126 (62.1%). Additionally, 110 (54.2%) respondents had received snakebite training, and 116 (57.1%) mostly lived in rural areas. The healthcare workers reported that farmers 130 (64.0%) were mainly bitten during the rainy season 114 (56.2%), in their farms or bush 129 (63.5%), and between 9 am to 12 noon 119 (58.6%).

**Table 2.**
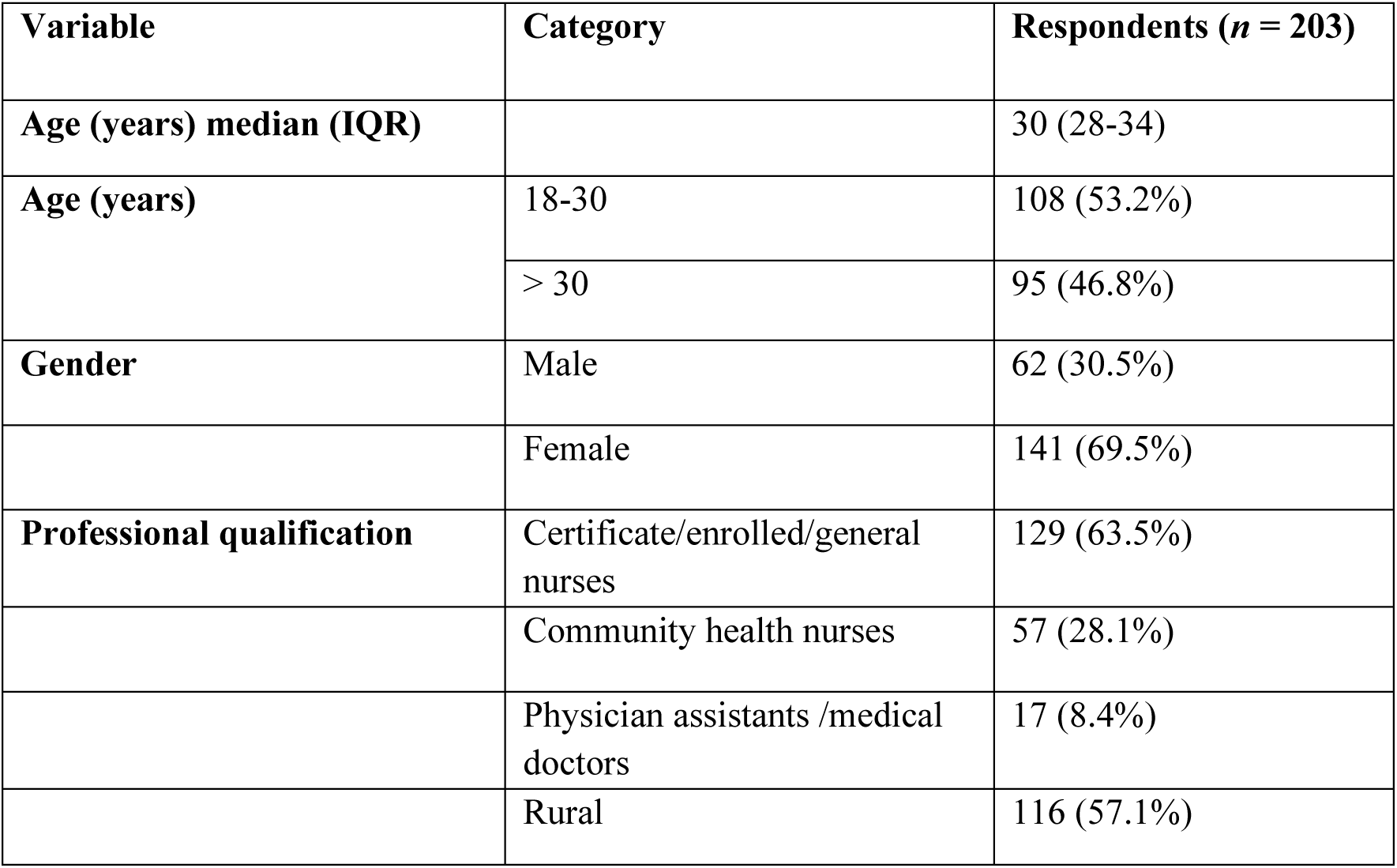
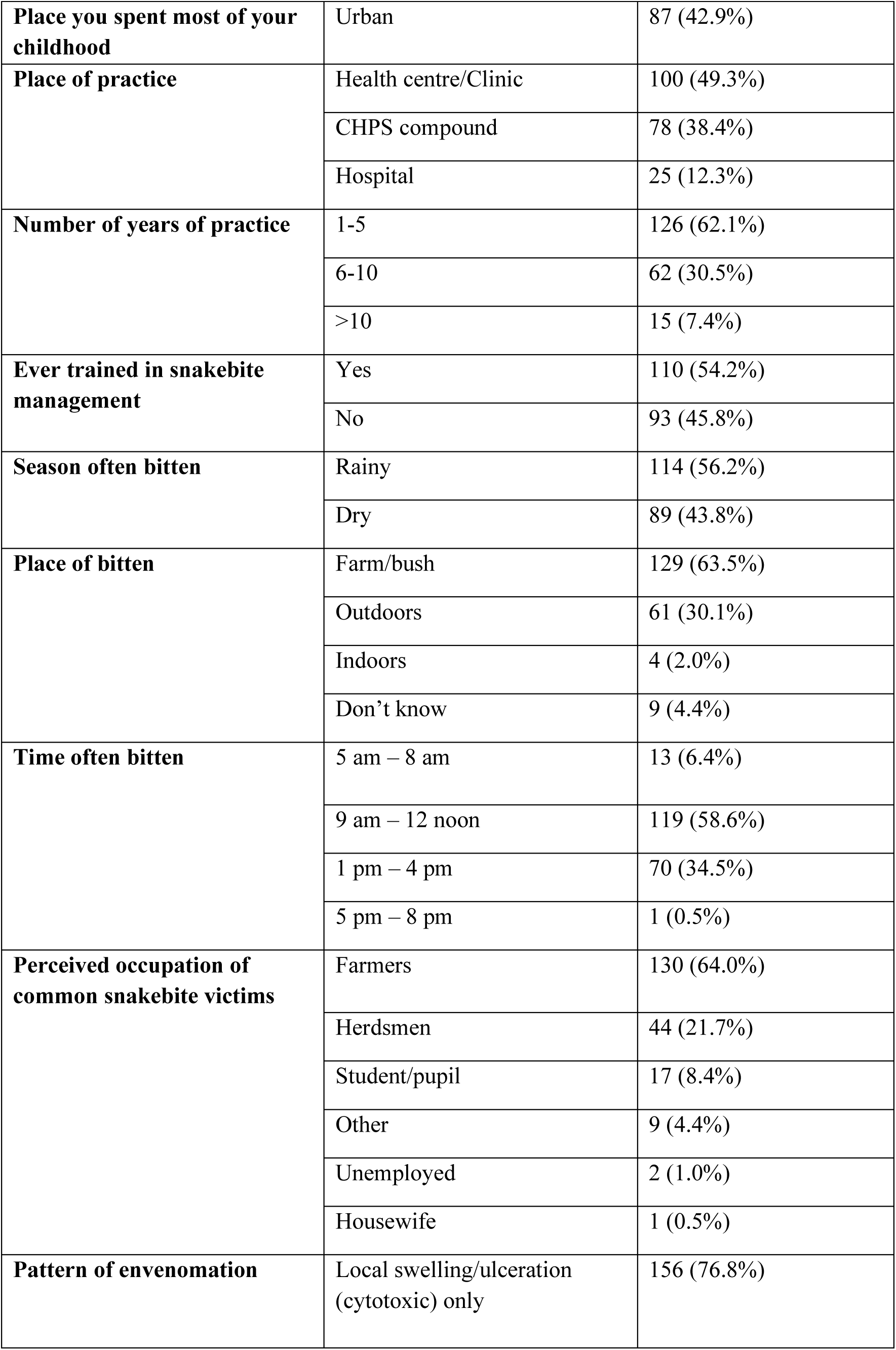
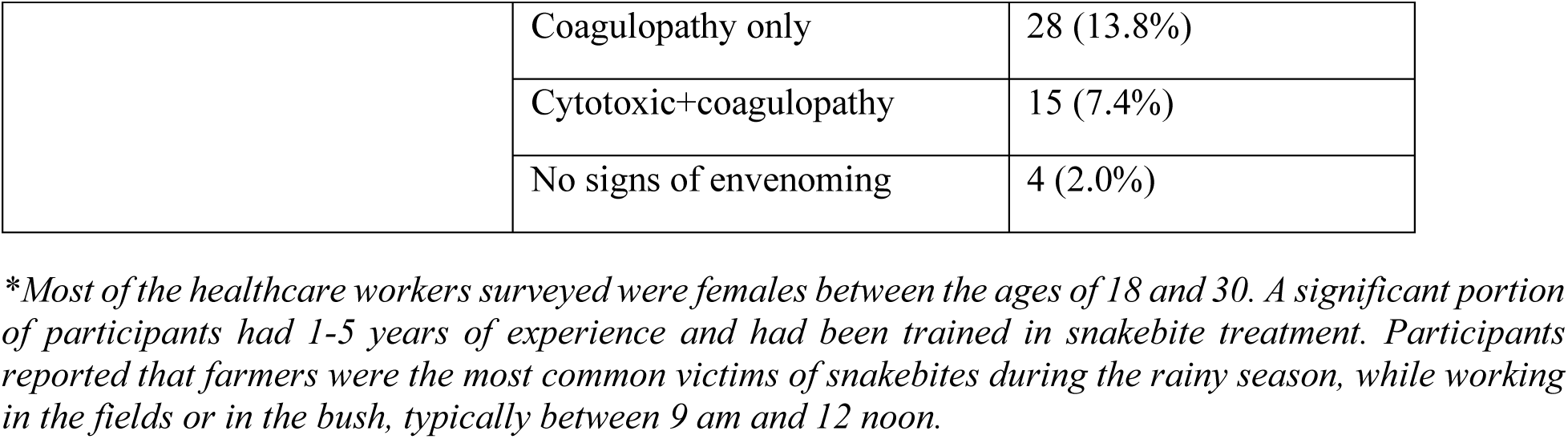
Demographic description of the study population.

Table 3 presents the model results of the snake antivenom products available for use in SSA. The results include utility estimates (UE), standard errors (SE), and confidence intervals (CIs). Figure 2 shows the relative importance healthcare workers attach to the snake antivenom products, represented by the UE and length of bars indicating the marginal probability (MP) values. All UE had the expected sign and significance at 95% CIs except for the Snake Venom Antiserum - Central Africa (Premium-CA) antivenom, which was insignificant. Among the antivenoms available for use in sub-Saharan Africa, participants highly prioritize Inoserp Pan-Africa (UE: 2.510; MP: 0.339; 95% C.I: 2.344, 2.681) and Snake Venom Antiserum-PanAfrica (UE: 1.548; MP: 0.129; 95%C.I: 1.419, 1.680). Other snake antivenoms with higher prioritization are EchiTabPlus (UE: 0.960; MP: 0.072; 95%C.I: 0.839, 1.082), followed by ASNA antivenom D (UE: 0.872; MP: 0.066; 95%C.I: 0.753, 0.992), ASNA antivenom C (UE: 0.799; MP: 0.061; 95%C.I: 0.682, 0.917), Snake Venom Antiserum African – 10 (UE: 0.668; MP: 0.053; 95%C.I: 0.552, 0.785), Anti Snake Venom Serum Pan Africa – 10 (UE: 0.658; MP: 0.053; 95%C.I: 0.542, 0.774), and Fav-Afrique (UE: 0.522; MP: 0.046; 95%C.I: 0.407, 0.637). These results suggest that some of the highly prioritized antivenom products may not necessarily be effective/quality but most commonly available, used, or familiar to the workers.

**Table 3.** Maximum difference model results of snake antivenoms available for treating snakebite envenoming.

| Snake antivenoms | Parameter estimates | SE | Lower 95% | Upper 95% |
| --- | --- | --- | --- | --- |
| Anti Snake Venom Serum Central Africa -6 | -0.239 | 0.057 | -0.351 | -0.127 |
| Anti Snake Venom Serum Monovalent Echis ocellatus | -1.101 | 0.059 | -1.219 | -0.984 |
| Anti Snake Venom Serum Pan Africa - 10 | 0.658 | 0.059 | 0.542 | 0.774 |
| Antiserum -PanAfrica (Premium-A) | 1.548 | 0.066 | 1.419 | 1.680 |
| Anti-Snake Venom Serum Central Africa (VINS-CA) | -0.498 | 0.056 | -0.609 | -0.388 |
| Antivipmyn-Africa (Antivip-A) | -0.698 | 0.058 | -0.813 | -0.583 |
| ASNA antivenom C (ASNA-C) | 0.799 | 0.059 | 0.682 | 0.917 |
| ASNA antivenom D (ASNA-D) | 0.872 | 0.060 | 0.753 | 0.992 |
| Combipack of Snake Venom Antiserum (African-Ten) | -0.234 | 0.056 | -0.345 | -0.122 |
| EchiTABG | -1.330 | 0.061 | -1.452 | -1.209 |
| EchiTabPlus (ET-Plus) | 0.960 | 0.061 | 0.839 | 1.082 |
| Fav-Afrique (FAV-A) | 0.522 | 0.058 | 0.407 | 0.637 |
| Inoserp Pan-Africa (Inoserp-P) | 2.510 | 0.085 | 2.344 | 2.681 |
| Menaven, Snake Venom Antitoxin | -0.986 | 0.059 | -1.102 | -0.870 |
| SAIMR Echis | -0.213 | 0.056 | -0.324 | -0.102 |
| Snake Venom Antiserum - Central Africa (Premium-CA) | -0.111 | 0.057 | -0.223 | 0.001 |
| Snake Venom Antiserum (Echiven) | -1.097 | 0.060 | -1.215 | -0.979 |
| Snake Venom Antiserum African - 10 (Afriven 10) | 0.668 | 0.059 | 0.552 | 0.785 |
| Snake venom antiserum Echis ocellatus (VINS-Echis) | -1.374 | 0.062 | -1.497 | -1.253 |
| **L-R Tests** |  |  |  |  |
| L-R Chi-Square | 4506.541 |  |  |  |
| DF | 19 |  |  |  |
| *P-Value* | <0.0001 |  |  |  |

\*SE, standard error; L-R, likelihood ratio; DF, degree of freedom.

**Fig. 2.**
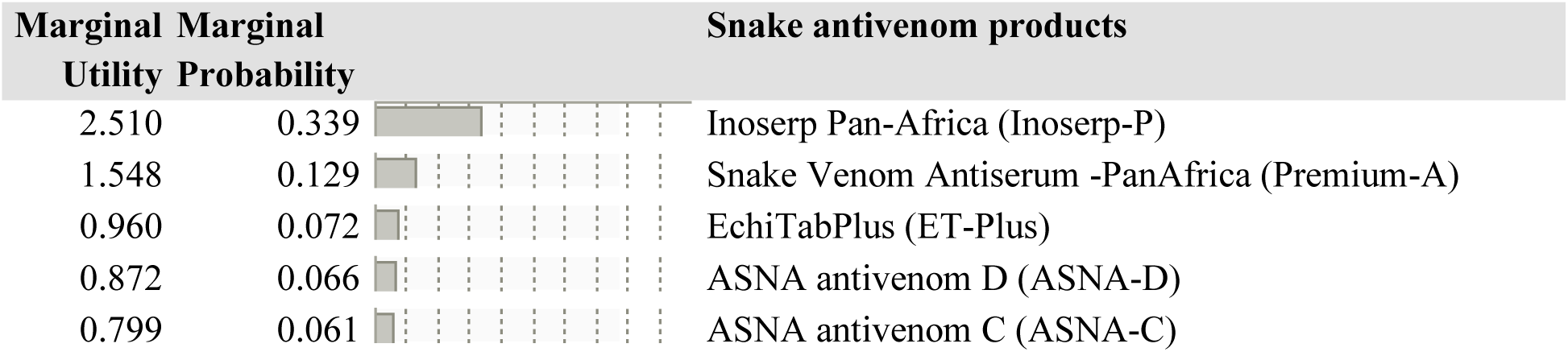
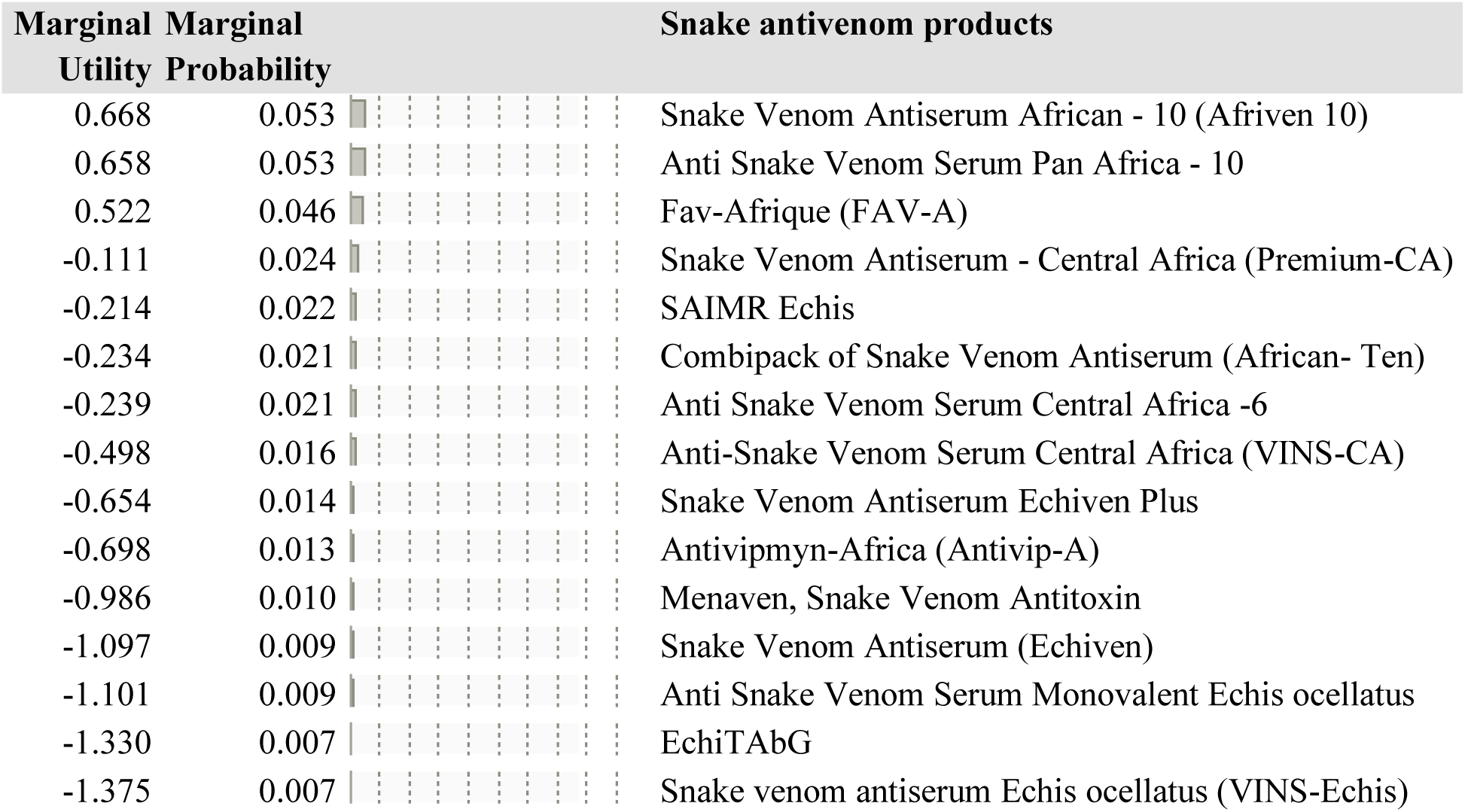
Marginal utility estimates and marginal probability of snake antivenoms available for treating snakebite envenoming. The corresponding marginal utility and probability values determine the importance attached to each snake antivenom.

Although significant, healthcare workers tend to trade-off or least prioritize some snake antivenoms, including SAIMR Echis, Combipack of Snake Venom Antiserum, Anti Snake Venom Serum Central Africa -6, Anti-Snake Venom Serum Central Africa, Snake Venom Antiserum Echiven Plus, Antivipmyn-Africa, Menaven, Snake Venom Antitoxin, Snake Venom Antiserum, Anti Snake Venom Serum Monovalent Echis ocellatus, EchiTAbG, as well as Snake venom antiserum Echis ocellatus (VINS-Echis) (UE: -0.214 to - 1.375; MP: 0.022 to 0.007; 95%C.I: -0.324, -0.102 to -1.497, -1.253).

Our results indicate that healthcare workers prioritize polyvalent and monovalent snake antivenom products differently. Some of the least prioritized antivenom products may not be commonly available, used, or familiar to these workers.

In Table 4 (in Appendix), we compare different snake antivenoms available for use in SSA with their associated marginal probabilities (MP), odds, and the greatest utility difference (GUD) between the antivenom products. Our findings revealed that there is a higher likelihood for healthcare workers to prioritize Inoserp-P antivenom (Odds: 48.66; MP: 0.979; GUD: 3.884; p=9e-227) compared to Snake Venom Antiserum Echis ocellatus (Odds: 0.020; MP: 0.020). Additionally, healthcare workers are more likely to prioritize Inoserp-P antivenom (Odds: 36.856 to 6.306; MP: 0.973 to 0.863; GUD: 3.607 to 1.841; p=2e-204 to 1.7e-65) compared to other snake antivenom products such as Snake Venom Antiserum African - 10 (Afriven 10) and Snake Venom Antitoxin (Odds: 0.027 to 0.158; MP: 0.026 to 0.136). On the other hand, the likelihood of healthcare workers prioritizing Snake Venom Antiserum -PanAfrica (Premium-A) (Odds: 0.382; MP: 0.276; GUD: -0.961; p=9.9e-20) is lower compared to Inoserp-P antivenom (Odds: 2.615; MP: 0.723).

However, participants are more likely to prioritize Snake Venom Antiserum -PanAfrica (Odds: 18.603 to 1.800; MP: 0.948 to 0.642; GUD: 2.923 to 0.588; p=7e-180 to 6.9e-11) compared to other snake antivenom products such as EchiTabPlus and Anti-Snake Venom Serum Central Africa (Odds: 0.053 to 0.555; MP: 0.051 to 0.357). Finally, there is a higher likelihood for healthcare workers to prioritize EchiTabPlus (Odds: 10.330; MP: 0.911; GUD: 2.335; p=5e-133) compared to other antivenom products such as Anti Snake Venom Serum Pan Africa – 10 and Fav-Afrique (Odds: 9.458 to 6.666; MP: 0.904 to 0.869; GUD: 2.246 to 1.897; p=6e-124 to 3.4e-95).

## 5. Discussion

Antivenom is a vital emergency medication used to treat snakebites, including pregnant women and people of all ages. Although patients receive antivenom treatment, healthcare workers are the primary end-users. As such, their antivenom prioritization is crucial to the research agenda and efforts to reduce the global burden of SBE. This study aimed to determine healthcare workers’ antivenom prioritization using the best-worst scaling methodology. The study provides quantitative evidence which could guide policy discussions on available snake antivenom products to enhance the procurement and supply of quality antivenoms, improve the efficacy of snakebite treatment, and elevate patient safety and care quality in health facilities in Ghana and SSA, where snakebite envenoming is a significant concern.

This study involved 203 healthcare workers, including community health nurses, physician assistants, medical doctors, and certificate/enrolled/general nurses. A prior study in Ghana indicated that medical doctors, registered nurses, physician assistants, community health nurses, and pharmacy technicians were the primary healthcare professionals involved in managing snakebites. In contrast, registered midwives and pharmacists were found to be the least engaged in snakebite management [29]. The study revealed that most participating healthcare workers were women aged between 18 and 30. Additionally, most respondents had received training in snakebite treatment. The healthcare workers reported that farmers were the most frequent victims of snakebites during the rainy season, often while working in their fields or the bush, typically between 9 AM and 12 noon. These findings are consistent with previous studies [47, 48, 49, 15, 14].

A key finding of this study was that healthcare workers’ highly prioritized Inoserp-P polyvalent snake antivenom. The other antivenoms that healthcare workers prioritized were Snake Venom Antiserum - PanAfrica, followed by ASNA-D, ASNA-C, Snake Venom Antiserum African - 10, Anti Snake Venom Serum Pan Africa – 10, and Fav-Afrique. However, some antivenoms were less commonly prioritized, such as SAIMR Echis, Combipack of Snake Venom Antiserum, Anti Snake Venom Serum Central Africa -6, Anti-Snake Venom Serum Central Africa, Snake Venom Antiserum Echiven Plus, Antivipmyn-Africa, Menaven, Snake Venom Antitoxin, Snake Venom Antiserum (Echiven), Anti Snake Venom Serum Monovalent Echis ocellatus, EchiTAbG, and Snake venom antiserum Echis ocellatus (VINS-Echis). It is worth noting that some of the highly prioritized antivenom products may not necessarily be effective/quality in treating SBE but most commonly available, used, or familiar to the workers.

Our results revealed that healthcare workers’ highly prioritized Inoserp Pan-Africa (Inoserp-P) antivenom. Earlier research reported that antivenom products with high market penetration [50, 21] were marketed without data on safety and clinical effectiveness from well-designed, pragmatic clinical trials [23], nor prior independent preclinical efficacy testing as well as poor preclinical efficacy [51, 52]. Therefore, multifaceted interventions are required, including but not limited to developing regional hubs for developing, evaluating, and regulating antivenoms alongside other recommendations by the WHO Strategy for Prevention and Control of Snakebite Envenoming [6]. An excellent strategy is WHO’s recent report on Target Product Profiles [2], which describes the characteristics needed to produce good antivenoms for Africa, thereby supporting the efforts towards designing and producing new antivenoms that respond to the African countries’ needs [20]. To reshape the market and improve compliance, national regulatory authorities should only register or authorize the marketing of antivenoms in the presence of independent preclinical neutralization tests and well-designed, pragmatic clinical dose-finding and safety studies [22, 53].

This study further revealed that healthcare workers prioritize Snake Venom Antiserum -PanAfrica, ASNA-D, ASNA-C, Snake Venom Antiserum African - 10 (Afriven 10), Anti Snake Venom Serum Pan Africa – 10, and FAV-Afrique. A study [54] similarly reported that ASNA Antivenom C and FAV-Afrique antivenom [55] are the most commonly used treatments for snakebites in Ghana. It is important to note that most of these antivenoms are polyvalent and are recommended for patients in areas where the venoms of different snake species produce similar clinical effects and where the dead snake is not available for identification. Further, although monospecific antivenoms are recommended for treating SBE caused by a known species of snake, they were least prioritized or traded off in this study. This suggests that monospecific antivenoms may be less commonly used for snakebite treatment, scarcer and more expensive. Efforts are needed to maintain a sustainable supply of appropriate antivenoms at a reasonable price [4, 2]. It is also important to implement regulations for available antivenoms, provide ongoing education and training to healthcare workers. Our study found that healthcare workers were more likely to prioritize certain antivenoms, such as Inoserp Pan-Africa and EchiTabPlus, than others. This finding highlights the need for the Ministry of Health to provide antivenoms following the recommendation of WHO, improve the availability and accessibility of antivenoms [56] in health facilities, as well as to ensure the quality and regulation of the product in the open market. Investigating alternative treatments, including immunomodulatory therapies from traditional medicines [10, 11], may offer complementary strategies alongside antivenom administration, potentially influencing healthcare workers’ priorities.

According to our findings, healthcare workers were more likely to prioritize Inoserp Pan-Africa (Inoserp-P) antivenom than snake venom antiserum Echis ocellatus (VINS-Echis). There is a lower chance of prioritizing Snake Venom Antiserum -PanAfrica (Premium-A) compared to Inoserp-P, but it is still more likely than other antivenom options. EchiTabPlus (ET-Plus) is more likely to be prioritized than some other antivenom options. Studies indicated that the choice of antivenom significantly influences certain treatment outcomes [57, 58, 59]. In contrast, other studies reported no significant impact [60, 61, 62]. Our results provide insight into the need to implement regulations for available antivenoms to improve patient safety and effective treatment. The application of computational models and bioinformatics tools [63, 64] has the potential to improve our understanding of venom components and antivenom efficacy. This may, in turn, influence healthcare workers’ preferences by informing them of the most effective treatments.

The authors acknowledge several limitations. Since the research was conducted solely in two districts of the Eastern Region, the findings may not accurately represent antivenom product prioritization by healthcare workers in other areas of Ghana and SSA. Although most respondents reported having received training on snakebite management, and the WHO recommends antivenom products that meet safety standards for deployment in primary healthcare facilities that have health workers who have been trained in the diagnosis and emergency treatment of snakebite envenoming [2], some healthcare workers interviewed may not regularly administer antivenoms. Additionally, even though they are experienced and had ever been involved in the management of snakebite victims in their facilities, some might have never administered antivenoms or possess varying levels of knowledge on the subject based on their professional backgrounds (such as community health nurses, physician assistants, medical doctors, and enrolled or general nurses). This variation in experience and knowledge may skew the findings toward the perspectives of the dominant cadre. Nevertheless, a prior study in Ghana indicated that medical doctors, registered nurses, physician assistants, community health nurses, and pharmacy technicians were the primary healthcare professionals involved in managing snakebites. In contrast, registered midwives and pharmacists were found to be the least engaged in snakebite management [29]. The antivenoms included in this study might not cover all available options, which could influence prioritization. Again, although the study investigated snake antivenom products available for treating SBE in SSA, some of the antivenoms may not be in the open market (Fav-Afrique, for example) or available for use in Ghana, including the Regional Medical Stores and health facilities. This may lead to biased utility estimates. Ultimately, further study is needed to confirm prioritization of actual antivenom products available for use in the Regional Medical Stores as well as in the various health facilities in Ghana, likely to be safe and efficacious, that its quality, safety, and efficacy are acceptable by WHO. There is also the need to assess whether these antivenoms are effective in local envenoming. Additionally, the study relied on provider reports or subjective judgments, and their integrity could not be verified. Therefore, detailed information from studies combining field and hospital data is needed to validate these results by reviewing hospital admission and discharge case report forms for administered antivenom products. There is an urgent need to assess antivenom availability and management in Ghana, including the public health system and medicine sales outlets such as pharmacies and licensed drug stores. Further study is needed to expand the classic health research toolkit by adapting existing robust methodologies like BWS and incorporating new tools such as artificial intelligence (AI), including machine learning (ML) and deep learning, to gather better snakebite information [65].

To conclude, this study presents crucial information to facilitate conversations on the antivenoms available in SSA for treating SBE. There is a pressing necessity to enforce regulations on snake antivenom products, improve procurement and supply of quality antivenoms, provide ongoing education, and offer training to healthcare workers to tackle the issue of SBE.

## Consent for publication

Not applicable

## Data availability statement

The datasets used and/analyzed in this study are available from the corresponding author upon reasonable request.

## CRediT authorship contribution statement

**Eric Nyarko:** Conceptualization, Methodology, Software, Validation, Formal analysis, Resources, Writing – original draft, Writing – review & editing, Visualization. **Ebenezer Kwesi Ameh:** Formal analysis, Resources, Investigation, Data curation, Project administration, Writing – review & editing.

## Acknowledgments

Eric Nyarko appreciates the fellowship support by the University of Ghana Building a New Generation of Academics in Africa (BANGA-Africa) project with funding from the Carnegie Corporation of New York [UG-BA/PD-003/2023]. We also extend our heartfelt thanks to all the informants who generously shared their knowledge and experiences.

## Appendix A. Supplementary material

**Table 4.**
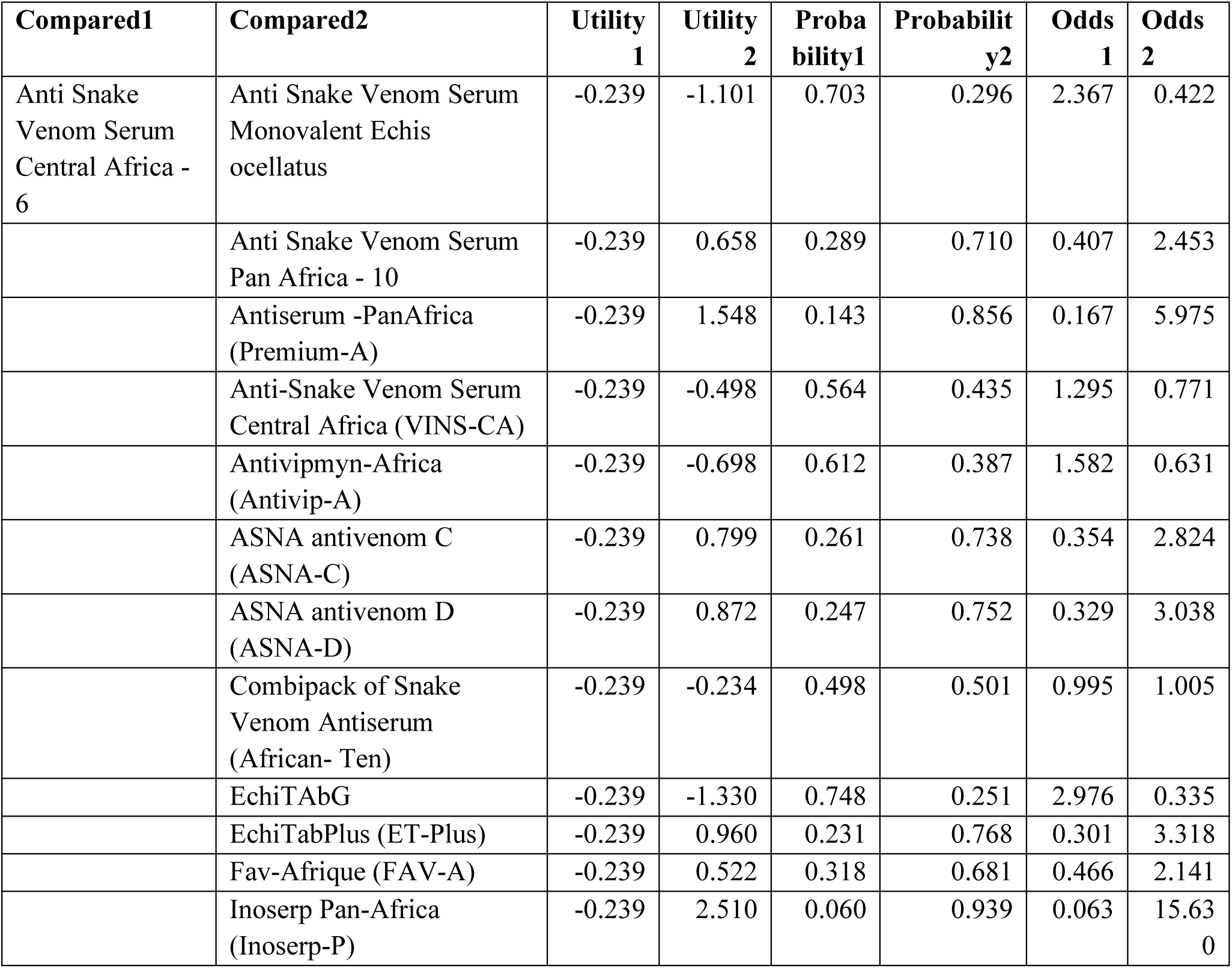
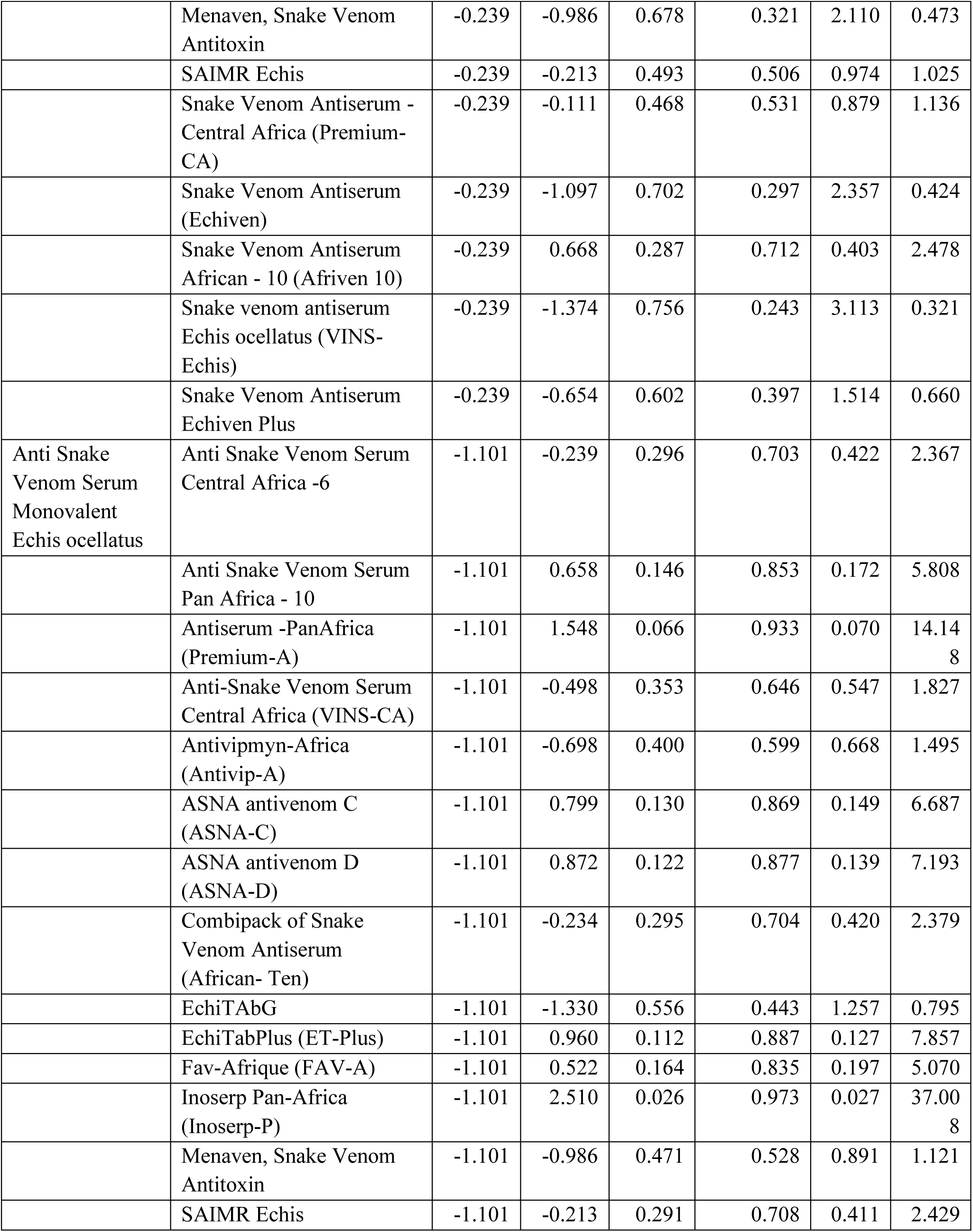
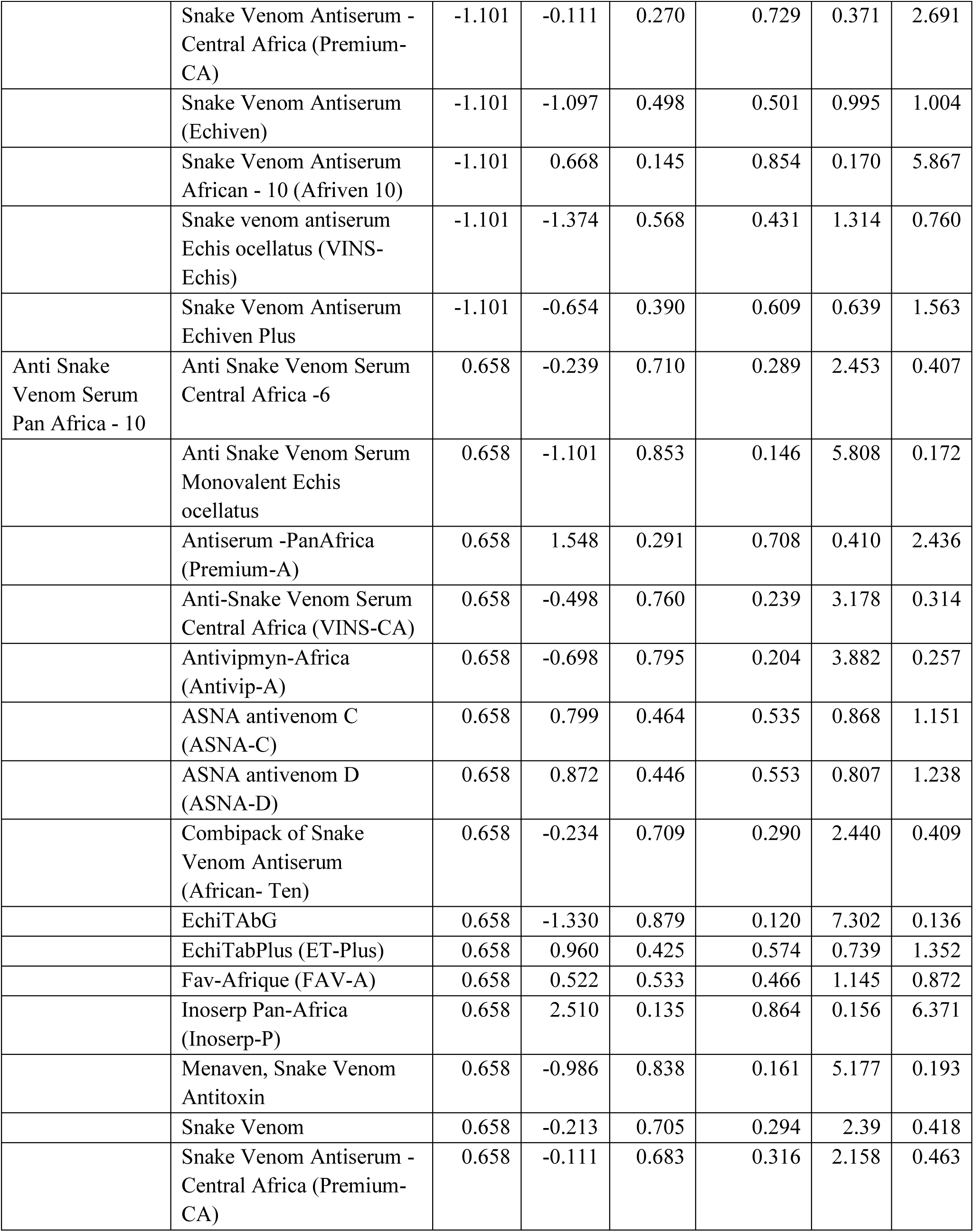
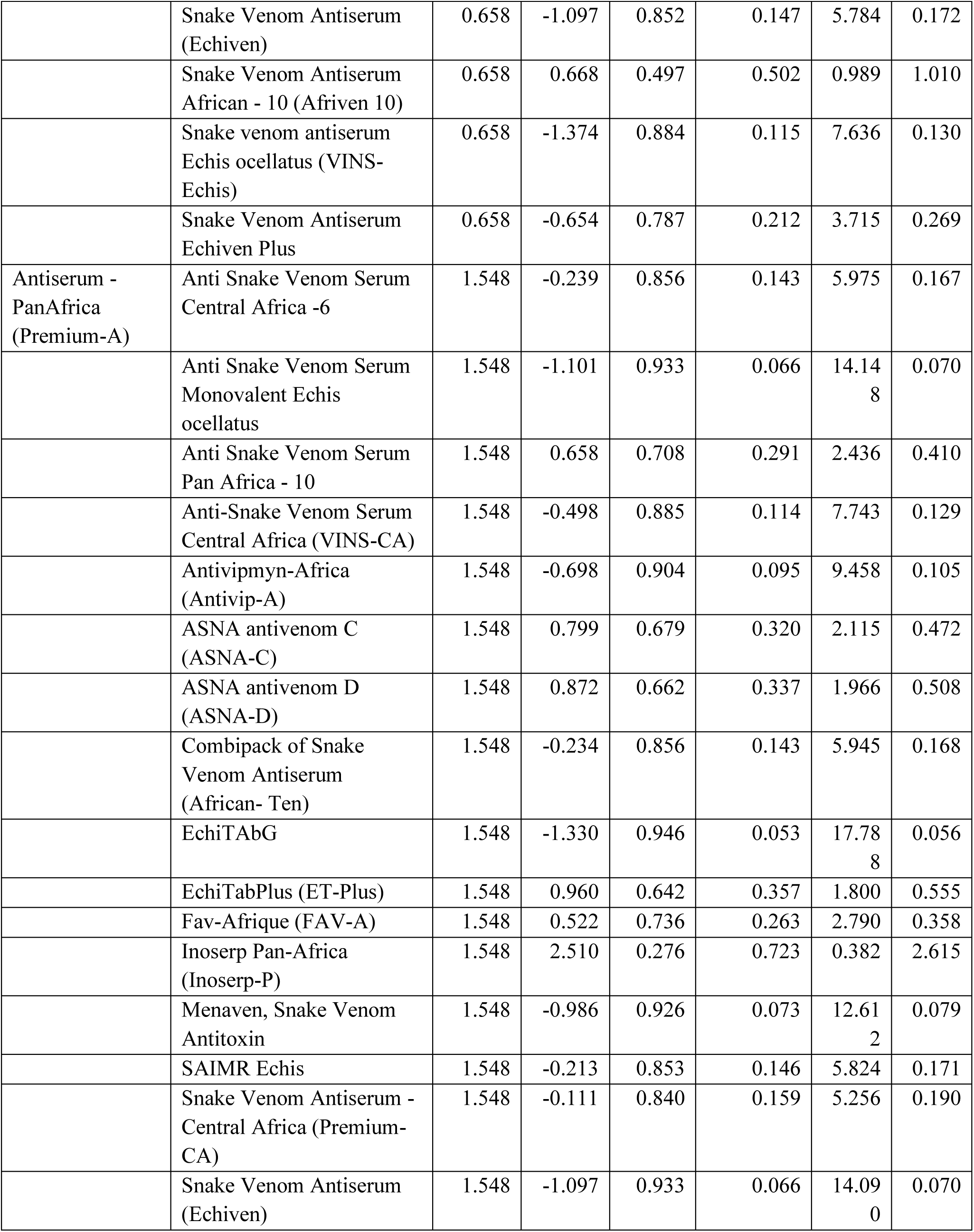
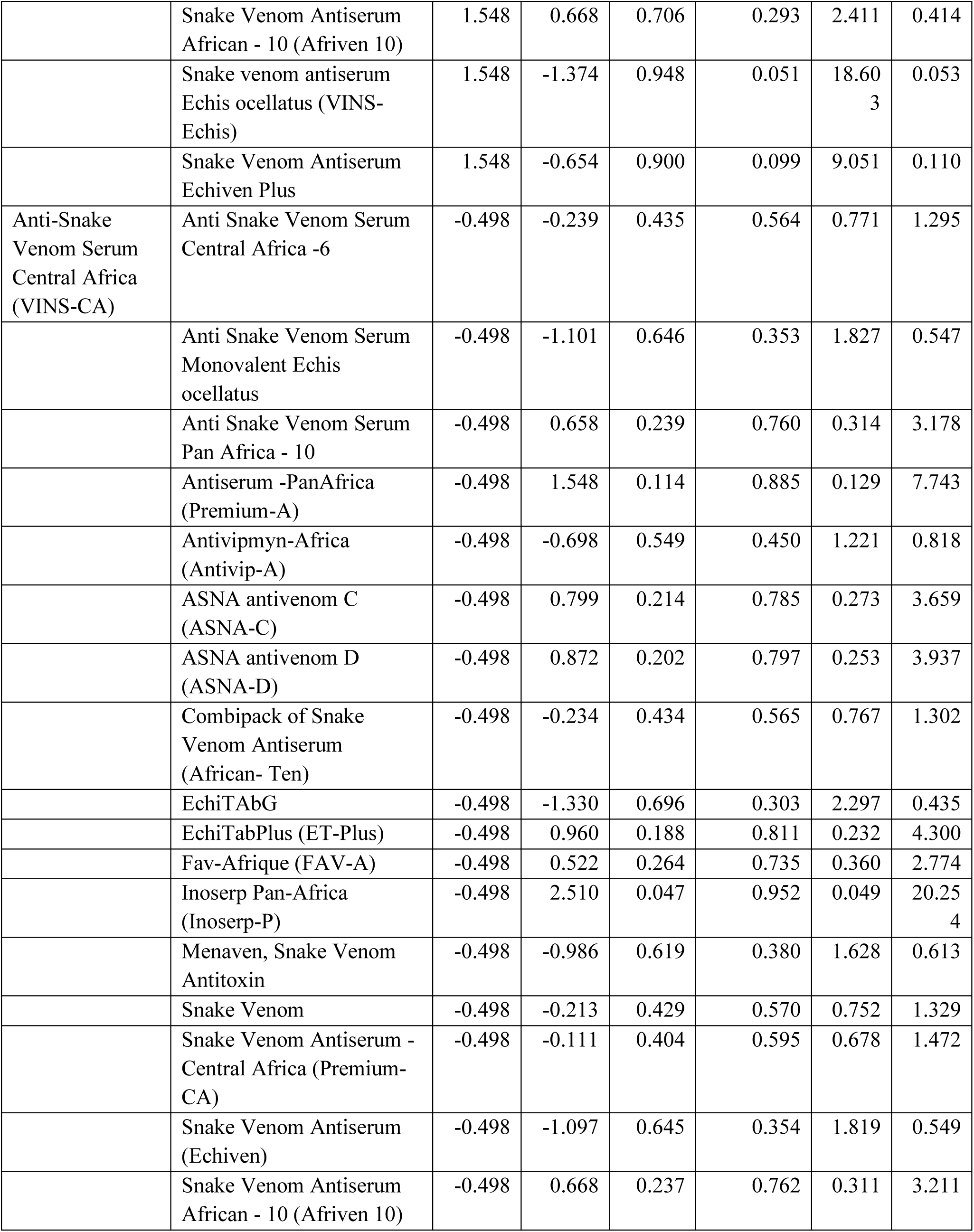
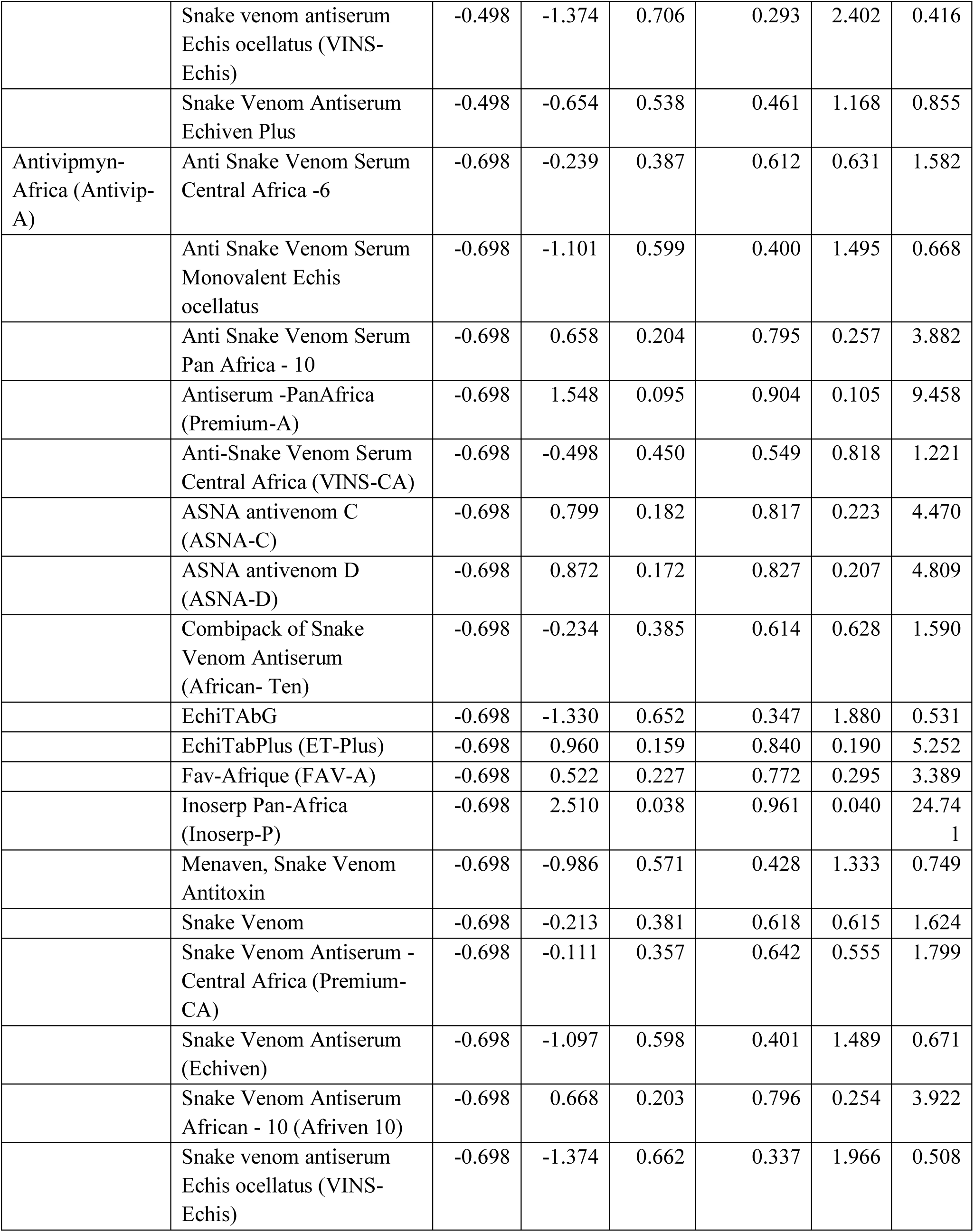
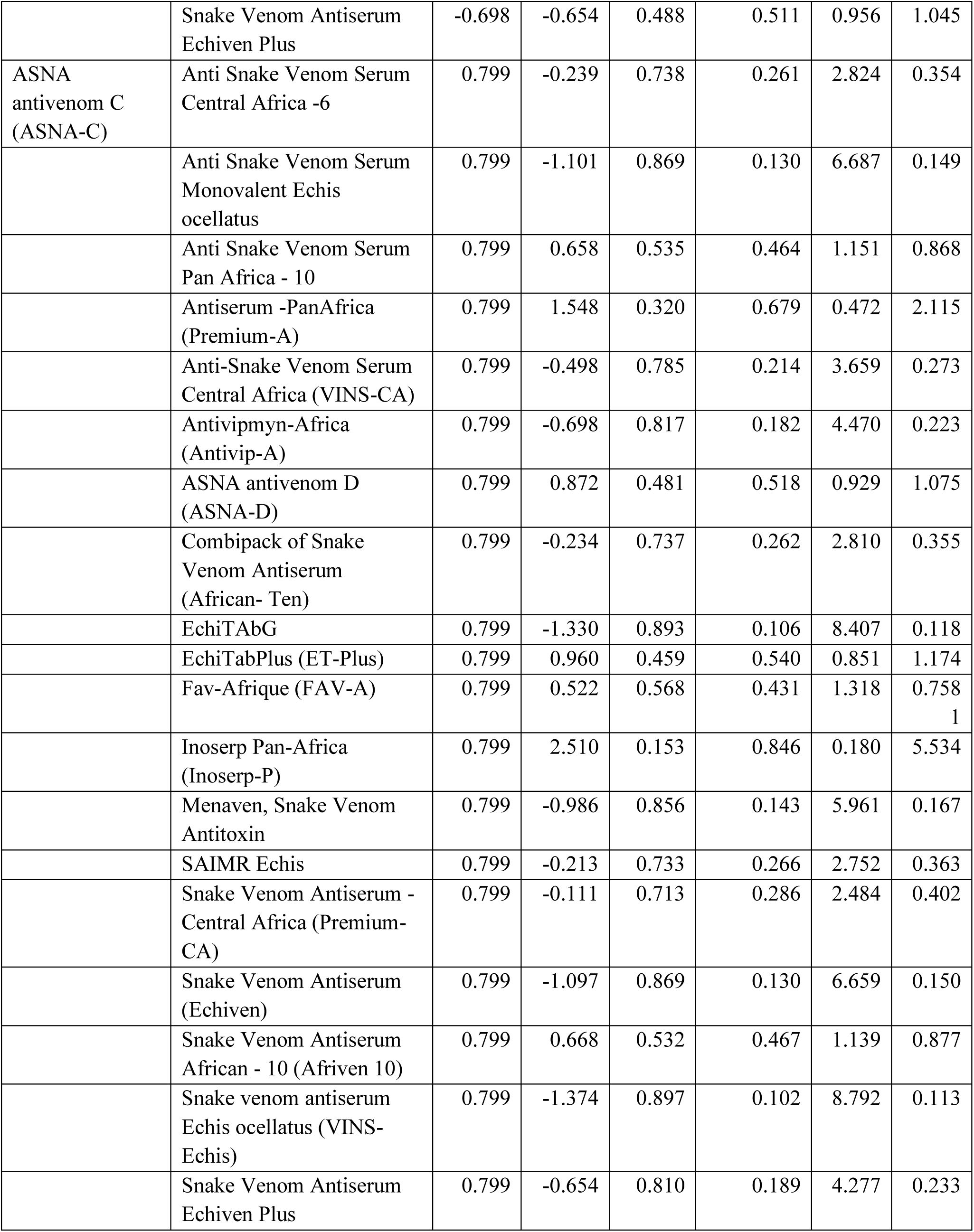
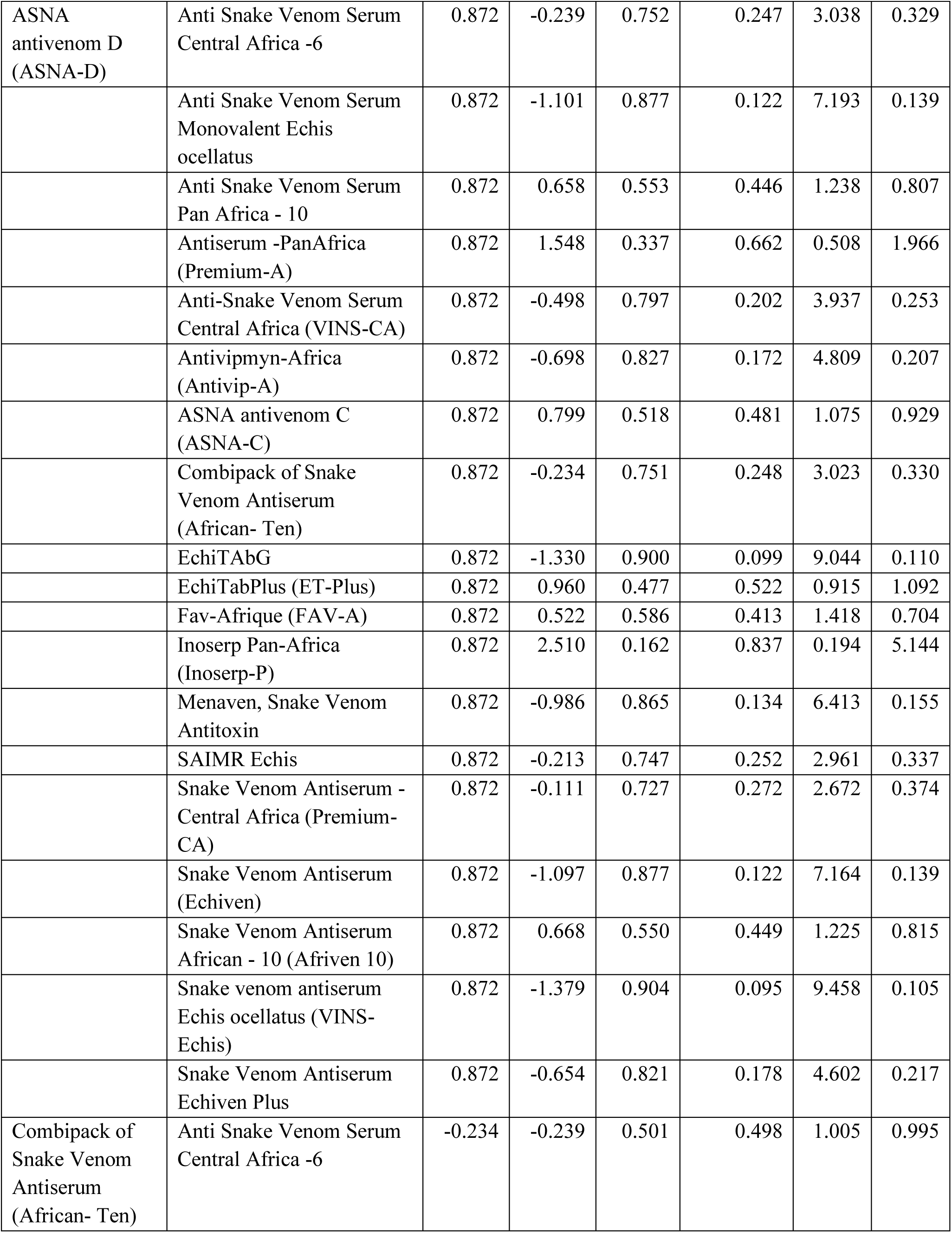
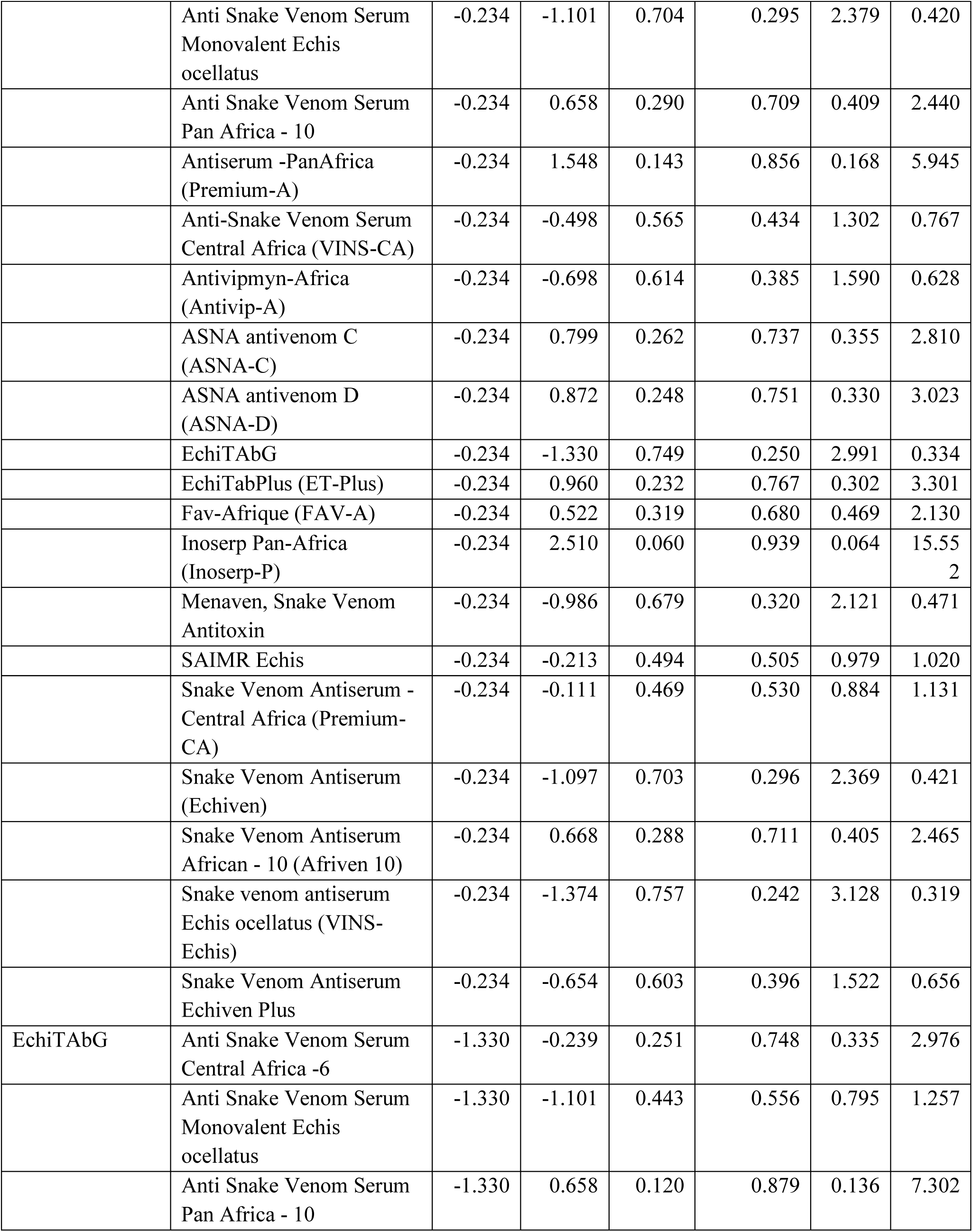
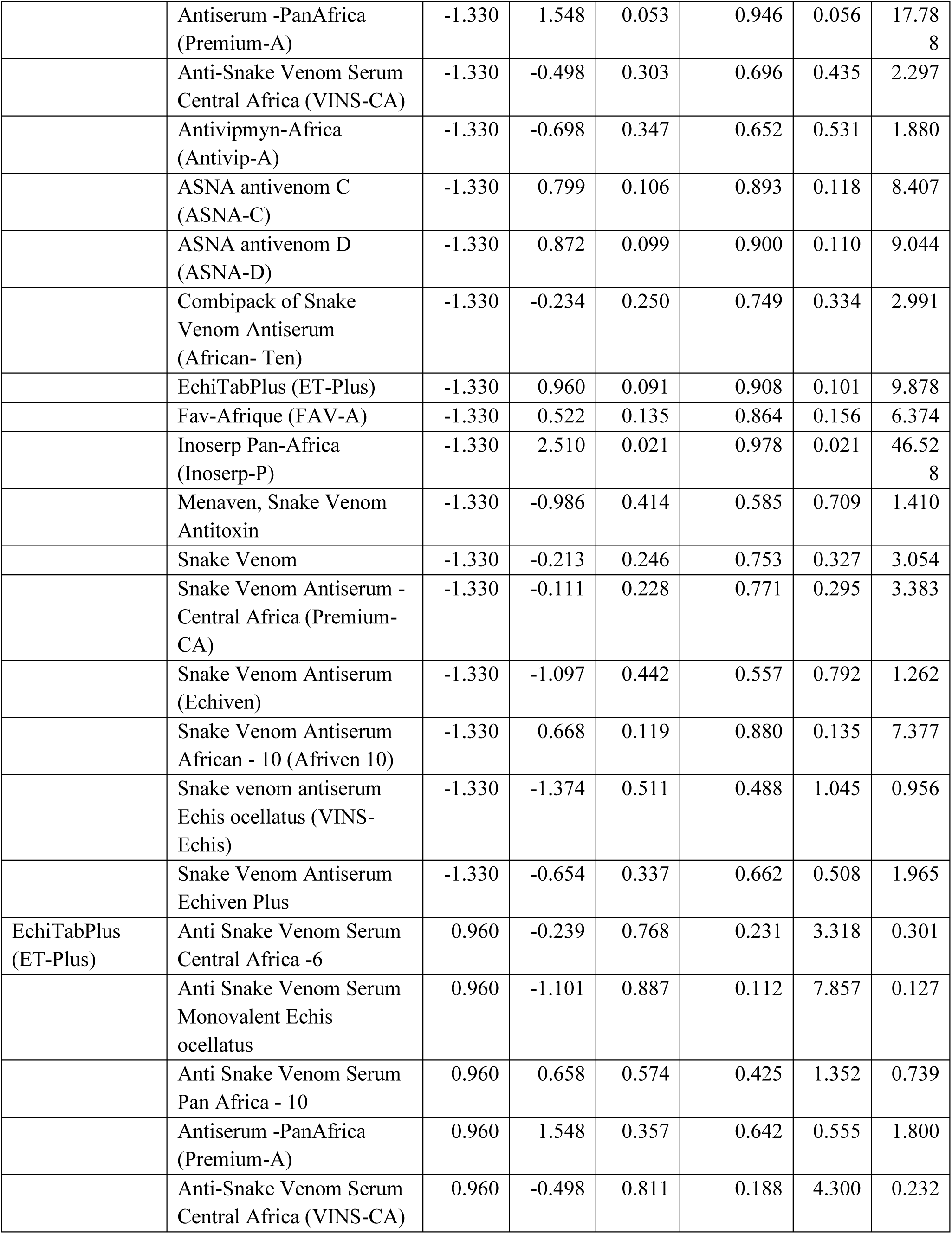
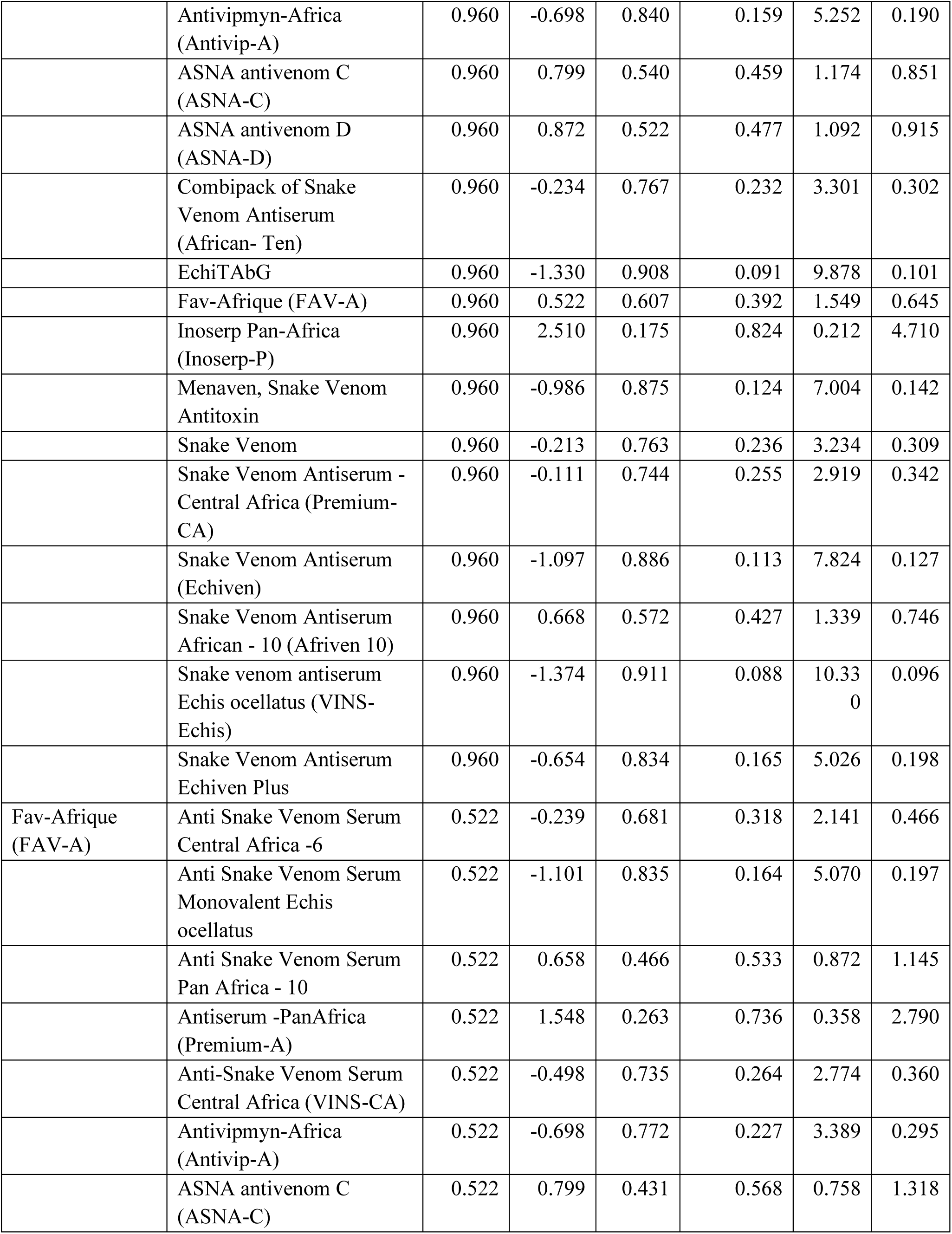
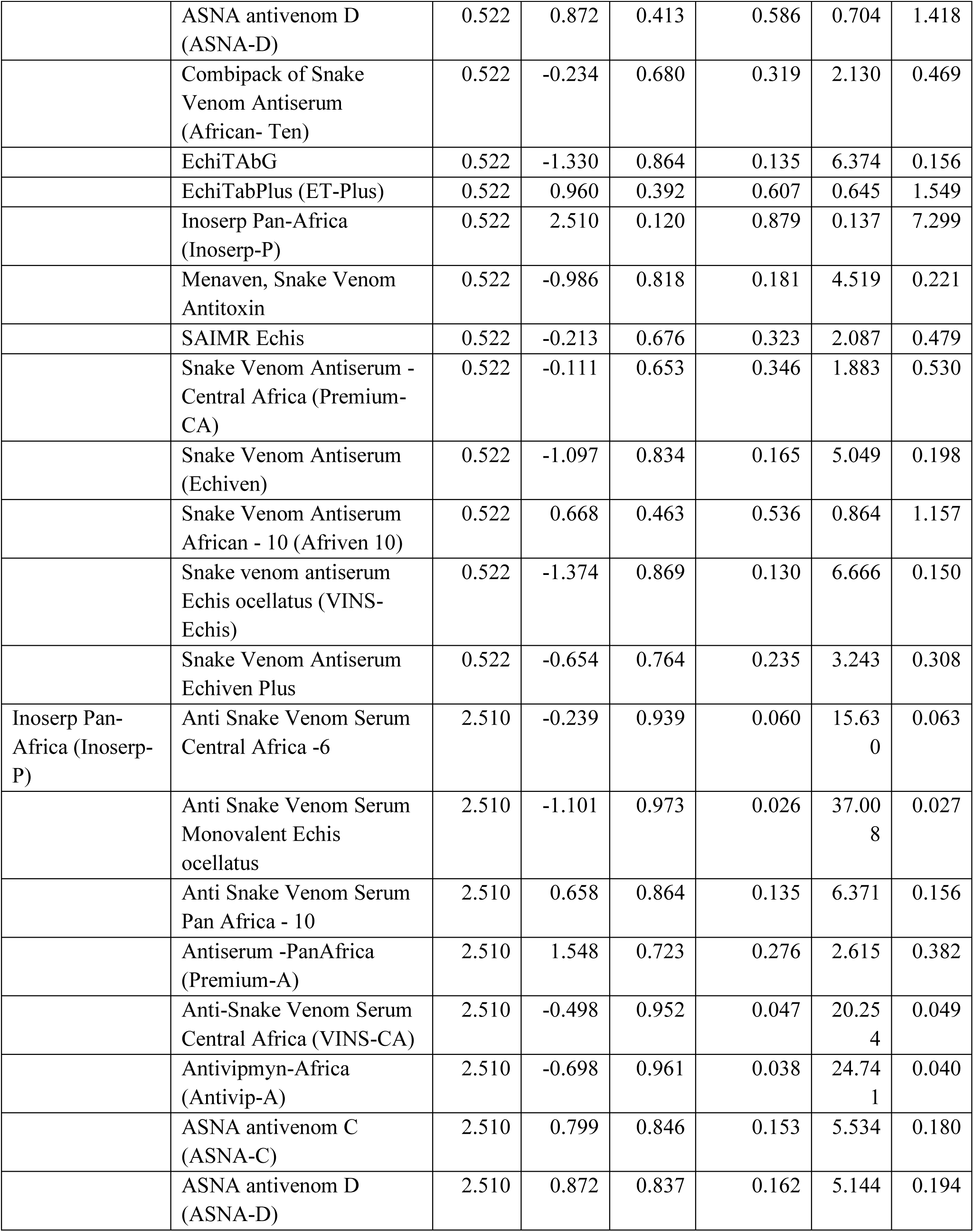
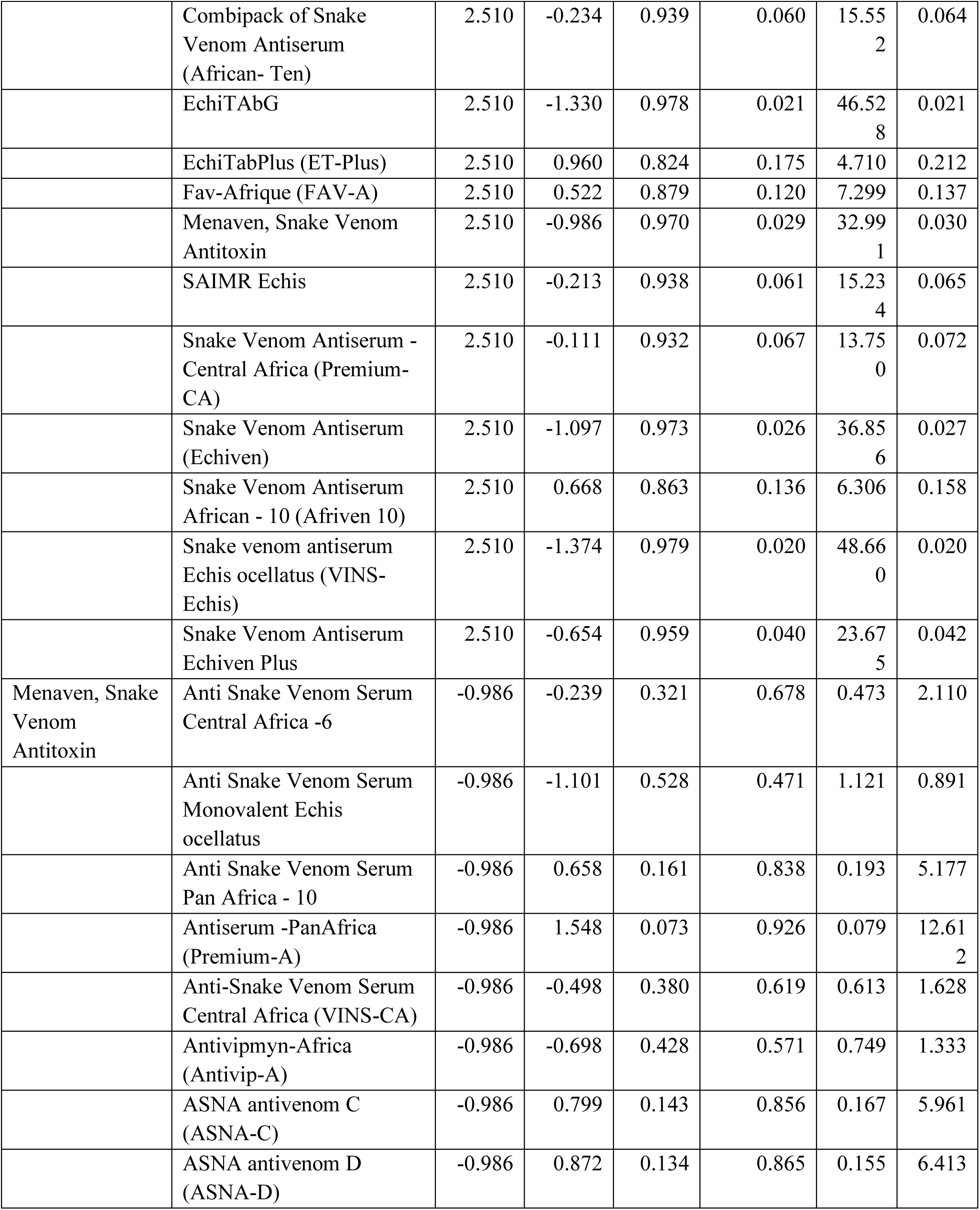
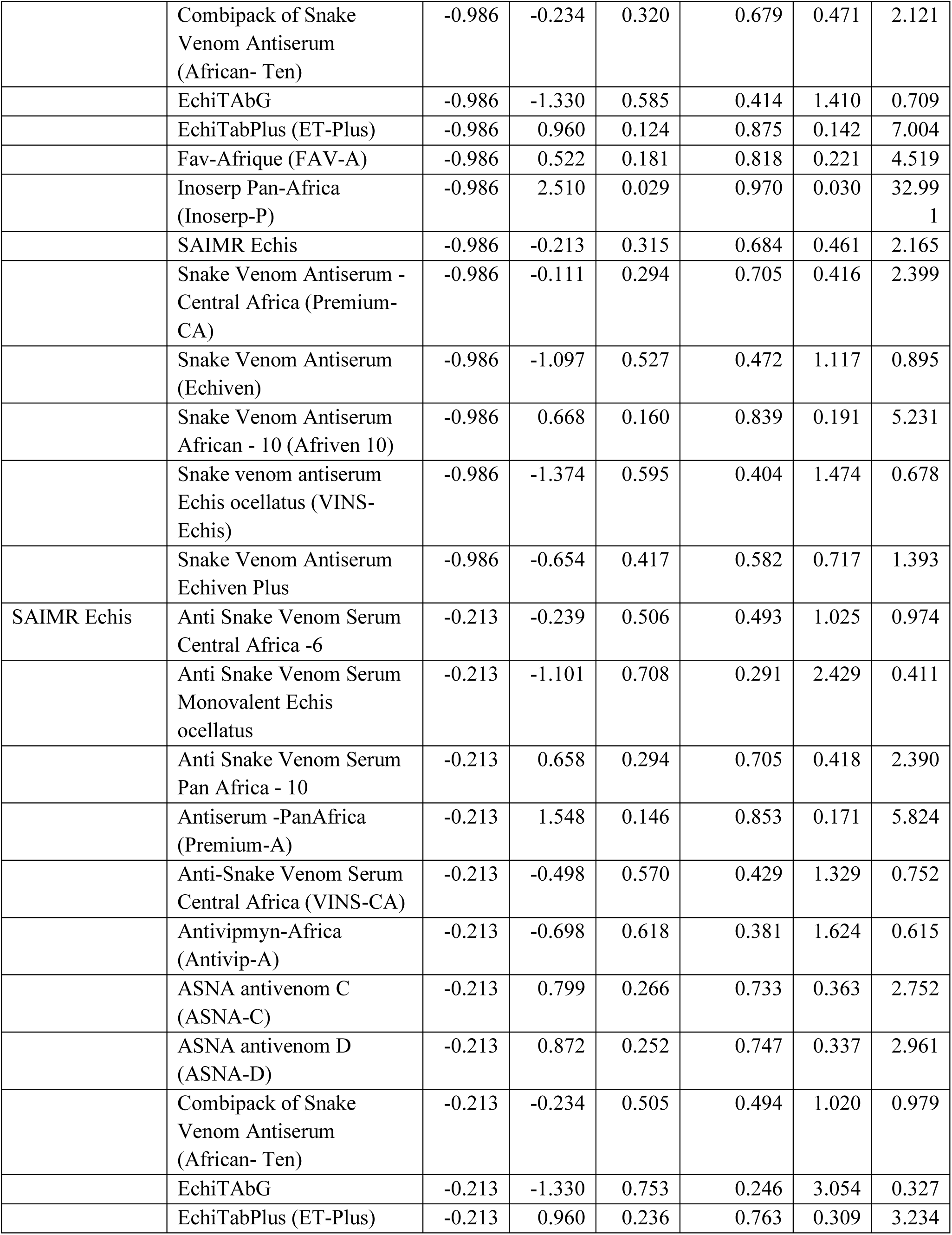
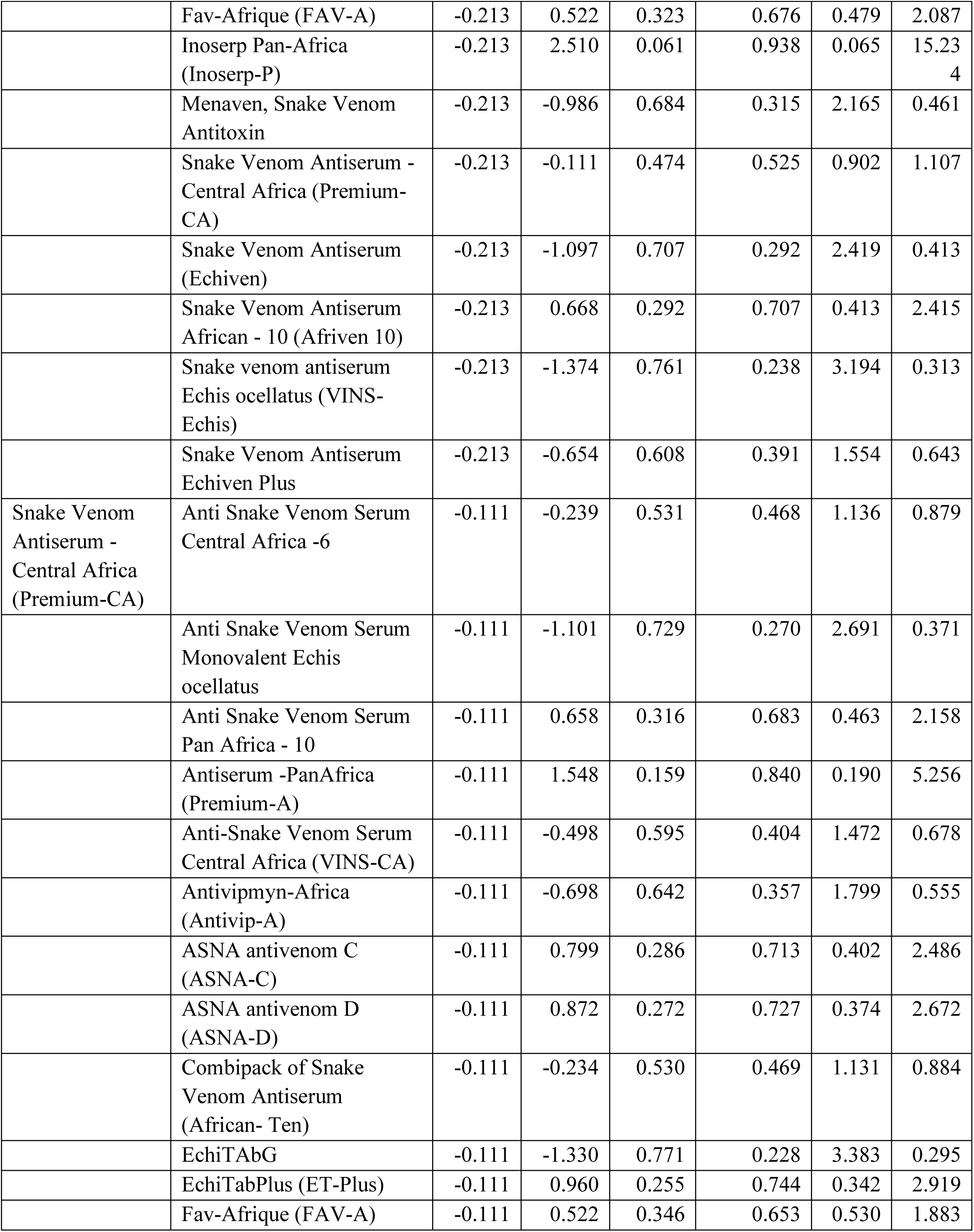
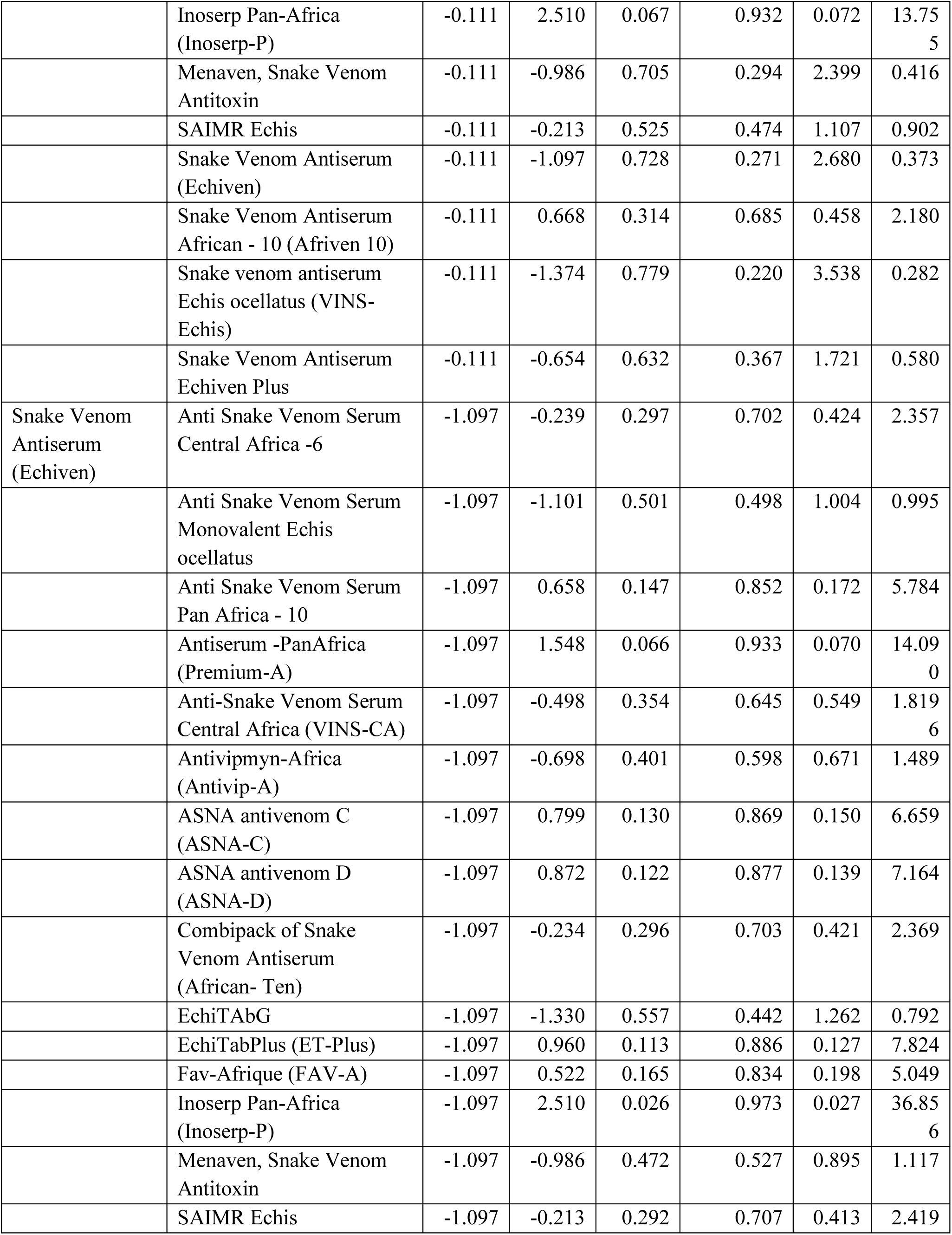
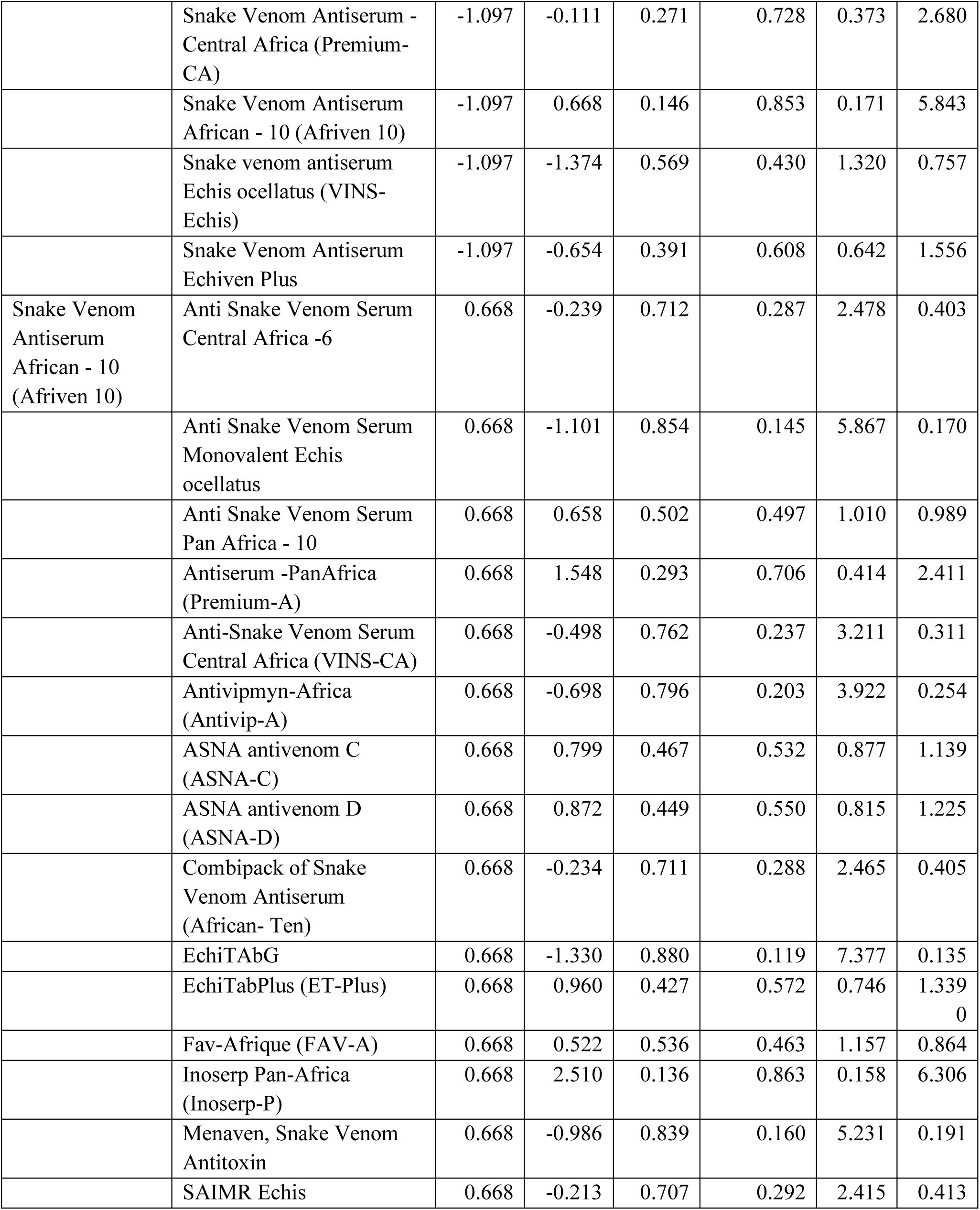
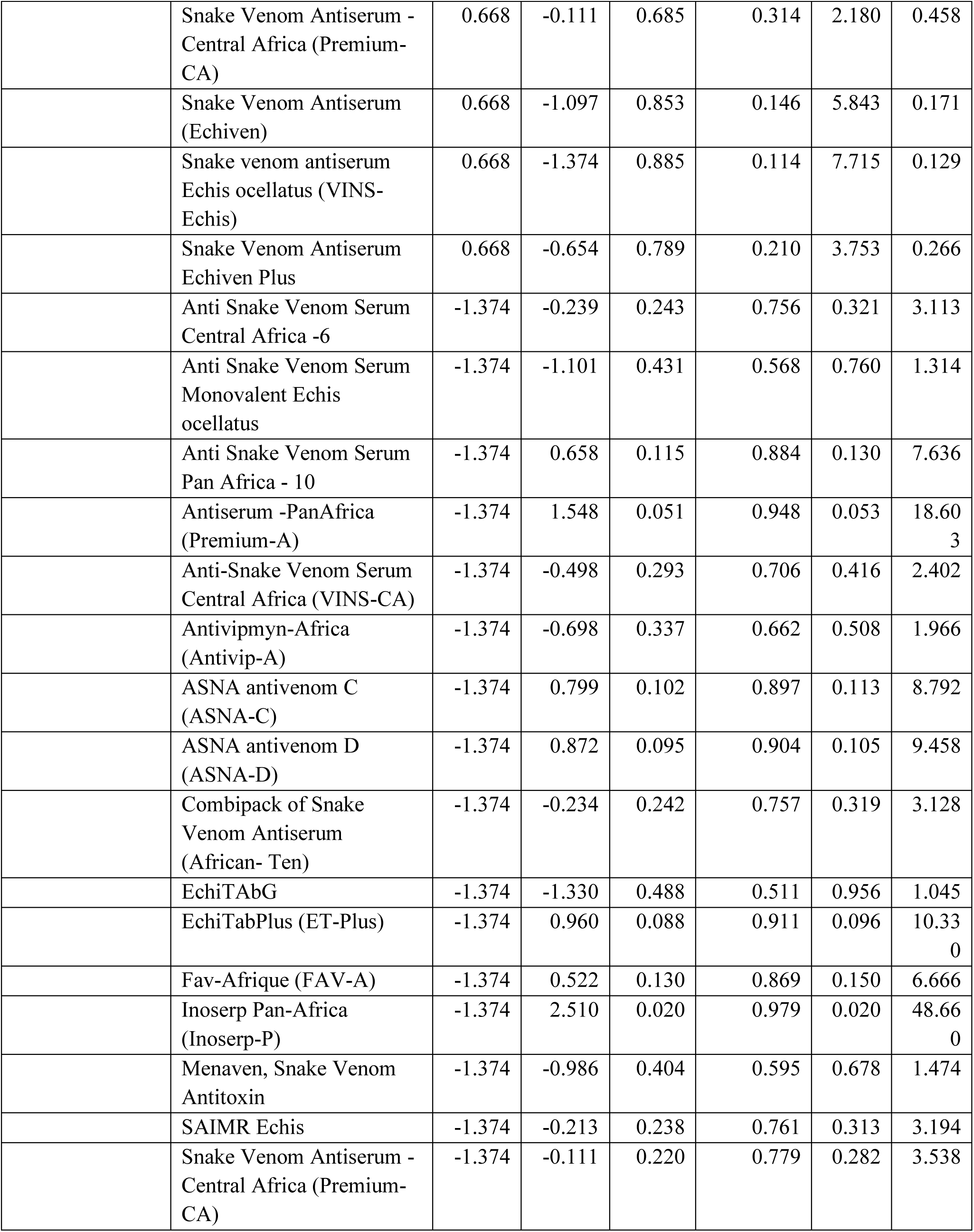
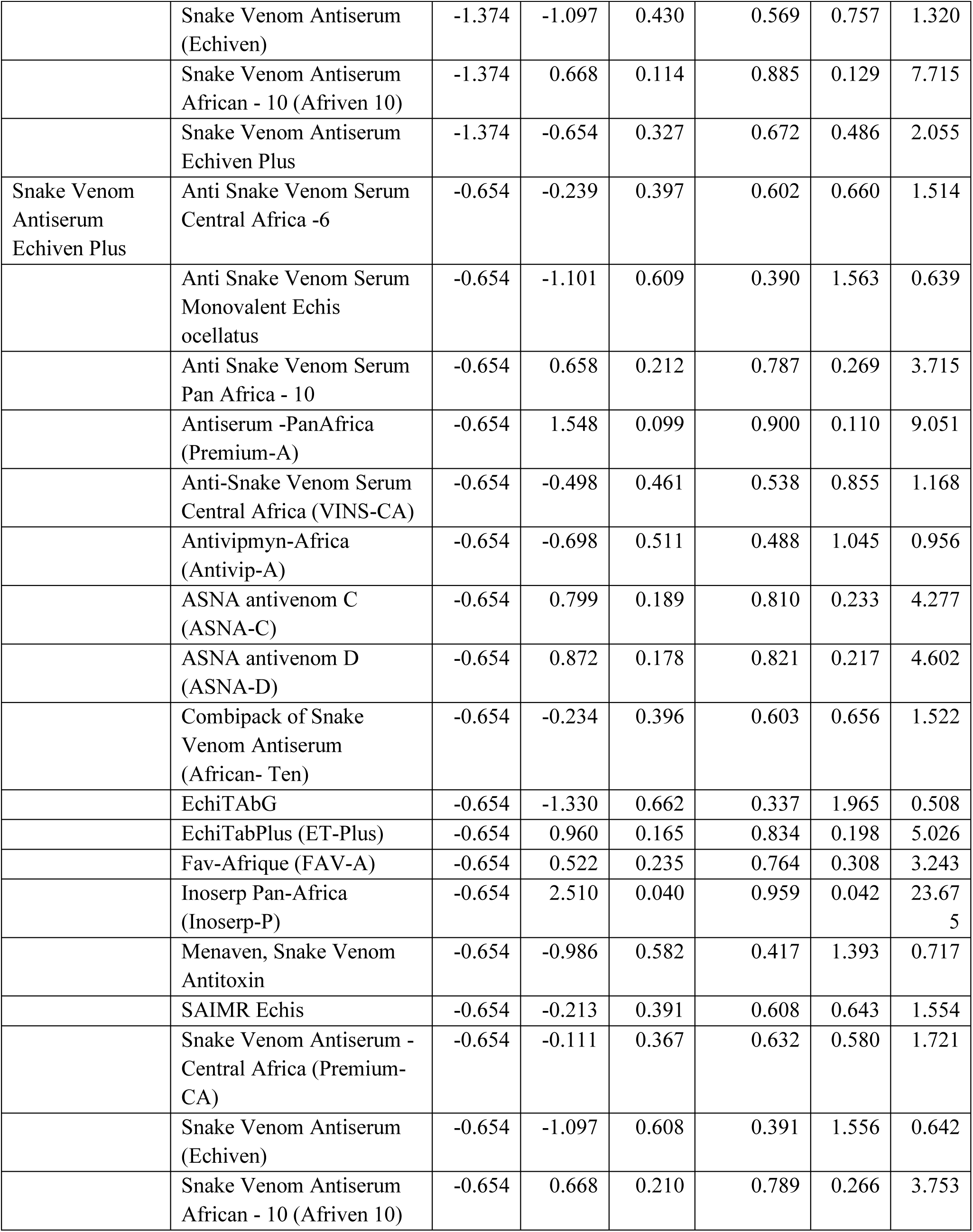
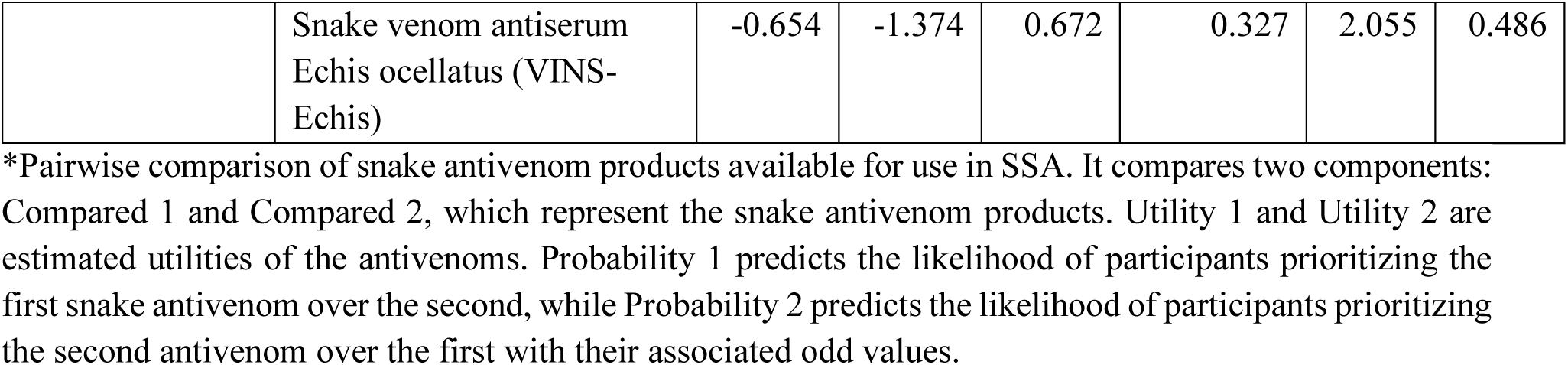
All levels comparison of snake antivenoms available for treating snakebite envenoming.

